# Validating the efficacy and value proposition of Mental Fitness Vocal Biomarkers in a psychiatric population: prospective cohort study

**DOI:** 10.1101/2023.11.21.23298774

**Authors:** Erik Larsen, Olivia Murton, Xinyu Song, Dale Joachim, Devon Watts, Flavio Kapczinski, Lindsey Venesky, Gerald Hurowitz

## Abstract

This study represents a practical advancement in the application of vocal biomarkers for mental health tracking in real-world settings. Through a prospective cohort study involving 104 participants from an outpatient psychiatric population, we introduced a novel “Mental Fitness Vocal Biomarker” (MFVB) score, derived from eight preselected vocal features supported by literature review. Our findings demonstrate the MFVB’s efficacy in objectively stratifying individuals based on risk for elevated mental health symptom severity using the M3 Checklist for transdiagnostic assessment (depression, anxiety, post-traumatic stress disorder, and bipolar) as reference standard. Continuous observation over time significantly improves efficacy, yielding a risk ratio of 1.53 (1.09-2.14, p=0.0138) for single 30-second voice samples to 2.00 (1.21-3.30, p=0.0068) for 2-week aggregations, depending on MFVB score. Notably, in the highly engaged subgroup (5-6 MFVB uses per week, 38% of participants), a risk ratio of 8.50 (2.31-31.25, p=0.0013) was observed, underscoring the utility of frequent and continuous observation. Participant feedback confirmed the user-friendliness of the application and perceived benefits, highlighting the MFVB’s potential as a cost-effective, scalable, and privacy-preserving adjunct to traditional psychiatric assessments. These results establish that vocal biomarkers are a promising tool for objective mental health tracking in real-world conditions, offering personalized insights into users’ mental well-being as they engage with clinical therapy or other beneficial activities that are associated with improved mental health risks and outcomes.

## INTRODUCTION

Associations between psychiatric conditions and alterations in vocal and speech characteristics are known from clinical practice and the domain of speech research. This association has prompted researchers to explore methods that could quantify the presence or severity of such conditions using automated vocal analysis and modeling approaches. Among other digital biomarker approaches, vocal biomarkers have a number of intrinsic advantages: they don’t require specialized hardware or complex procedures, and are considered among the promising digital biomarkers in psychiatry (1,2). Moreover, “smart” personal devices equipped with microphones and signal processing capabilities (e.g., phones, speakers, TVs, vehicle cabins) offer convenient and scalable means of reaching large populations and could help people track their mental wellbeing in ways that go beyond simply logging moods and feelings (3,4).

While considerable effort has been devoted to vocal biomarker discovery and predictive modeling based on data obtained in case-control studies and research settings, comparatively less work has been done to demonstrate technology that can both accurately measure and inform users about relevant vocal characteristics related to mental wellbeing outside of such controlled settings and then incorporate these into an application that provides results in a way that is understandable and informative to users. Such work is of high importance, as many digital health tools have low engagement and retention rates and thus do not fulfill their potential value proposition. Negative attitudes towards apps and wearables, lack of measurement agreement with gold standards, and missing evidence around efficacy are common issues with digital health tools (5). The present study aims to address those shortcomings by showing that a vocal biomarker-based journaling application can provide meaningful results regarding mental wellbeing to users that is calibrated against validated assessment tools, in a way that engages and benefits users. We do this by validating previously identified vocal features relevant to a range of psychiatric conditions. We contrast our results with those obtained in similar studies in the Discussion section.

Vocal feature selection for such an application is complex due to the many potential reasons for vocal changes in individuals with psychiatric conditions. For example, anxiety or stress can increase muscle tension, including the muscles involved in vocal production and leading to issues such as throat strain, fatigue, or tightness (6,7). Anxiety-related behaviors, such as heightened vocal effort and frequent throat clearing, can contribute to voice problems (8). Chronic hyperventilation, common especially among people with panic disorders, can affect prosody and vocal strength. Individuals experiencing depression may exhibit reduced self-care practices, including inadequate hydration, which can result in vocal cord dryness and subsequent voice changes (9). Certain medications prescribed for psychiatric conditions may produce side effects that impact voice quality or prosody (10–13). Mental health disorders can coincide with dietary changes or sleep disturbances, potentially leading to vocal fatigue, hoarseness, or other vocal issues (14). Alterations in muscle control or tension can be caused by neurotransmitter imbalances or hormonal shifts associated with depression and anxiety (7). Chronic stress, a hallmark of many psychiatric conditions, can lead to chronic inflammation or frequent infections, including of the vocal cords (15). Changes in brain activity, connectivity or functioning may contribute to psychomotor retardation or reduced cognitive resources and thereby affect speech production (16,17). As part of the broader slowing down of physical and cognitive processes, psychomotor retardation may contribute to typical speech pattern changes in depression such as slowing of speech, increased pauses, and a flattening of prosody (18–22). Conversely, voice disorders may also create a predisposition for individuals to develop mental health issues. For instance, voice disorders can lead to social withdrawal, communication challenges, stigma, and an overall reduced quality of life. These consequences may give rise to feelings of loneliness, low self-esteem, stress, anxiety, and depression (23–25). Voice therapy can improve mental health symptoms, especially in high-risk populations (26). Reviews of specific vocal changes identified in different psychiatric conditions highlight this complexity (27,28).

Several previous studies have addressed the vocal feature selection issue using machine learning approaches, using vocal feature values as inputs, diagnosis (or symptom questionnaire responses) as training labels, and health prediction scores as outputs (29–39). While some of the studies have showed high levels of performance in differentiating populations with higher vs. lower mental symptoms (or diagnosed vs. healthy controls), most have not been validated on new datasets. Most studies were also retrospective case-control designs, where data was obtained from research participants in controlled settings that did not interact with a digital application that could provide them with feedback regarding their mental wellbeing.

To test the potential for vocal biomarkers in real-time mental health tracking, we conducted a prospective cohort study with a predesigned vocal biomarker application that provides users with direct feedback. Our vocal features selection was hypothesis-based, using accumulated evidence from published research, and visualize the results in a digital health app that study participants use on their personal smartphone. To simplify interpretation of results, these features are further aggregated into a single composite score which we call “Mental Fitness”. This terminology is intended to convey the non-medical nature of the score, avoid stigma associated with illness or disorders, and suggest an analogy with physical fitness – strength, resilience, and ability to rapidly recover from injury or setbacks. The “Mental Fitness” concept underscores the opportunity for individuals to engage in healthy behaviors that can proactively mitigate mental health risks, fostering well-being and mental resilience akin to how physical fitness promotes overall health and vitality.

In parallel to the success of tools that monitor and enhance physical activity, our focus on Mental Fitness aims to enhance mental self-awareness and foster proactive mental well-being. Much like the established benefits of activity self-monitoring for promoting physical health (40), we envision Mental Fitness tracking as a catalyst for fostering mental resilience. Providing users with this familiar fitness tracking framework could facilitate better understanding and improving of their mental well-being within an integrated digital ecosystem, extending the benefits seen in the physical health domain to empower individuals in actively managing their mental fitness.

We used the M3 Checklist, a clinically validated 27-item questionnaire that provides an overall mental health symptom severity result and by subdomains of depression, anxiety, post-traumatic stress disorder (PTSD), and bipolar disorder as standard for mental health symptoms (41,42). This provides an assessment of vocal analysis efficacy across mental health domains as well as an assessment of how total mental health symptom severity (regardless of type, i.e., transdiagnostically) impacts voice production. Vocal biomarker performance will be assessed by evaluating the relative risk and risk ratio for the presence of moderate or higher symptom severity in predefined vocal biomarker score ranges; value proposition will be assessed through user feedback and engagement with the study app. We tested this approach in a clinical population receiving outpatient treatment for common mental disorders. Study duration was 4 weeks to provide sufficient opportunities for participants to use the vocal biomarker study app and provide informed feedback. The study was conducted at a single site (Cognitive Behavioral Institute, Pittsburgh PA), although a pilot phase also included St Joseph’s Healthcare Hamilton (Hamilton, ON).

## METHODS

### Mental Fitness study app

Vocal biomarker analysis and visualization was accomplished using a smartphone app developed by Sonde Health, described in detail in the Supplemental Material. Briefly, the app implements a voice journaling capability where users can record 30 second voice samples in response to a range of predefined prompts or talk about their own thoughts and feelings. The response is automatically transcribed and stored in a journaling section, while the recorded audio is used for vocal biomarker analysis using methods similar to those described in OpenSMILE (43) and Praat (44). A “Mental Fitness” Vocal Biomarker (MFVB) score and its feature components are presented to the user after each journaling session and stored in a history section.

The MFVB score components were selected based on a literature review of well supported vocal features relevant to mental health, in particular depression. The features measure different systems involved in vocal production – briefly, they include jitter and shimmer (45–47), pitch variability (45,47–50), energy variability (45,51), vowel space (52,53), phonation duration (37), speech rate (29,50,47,54), and pause duration (50,29,55,47,56). More information on these features is provided in Table 1 and the Supplemental Material.

**Table 1.**
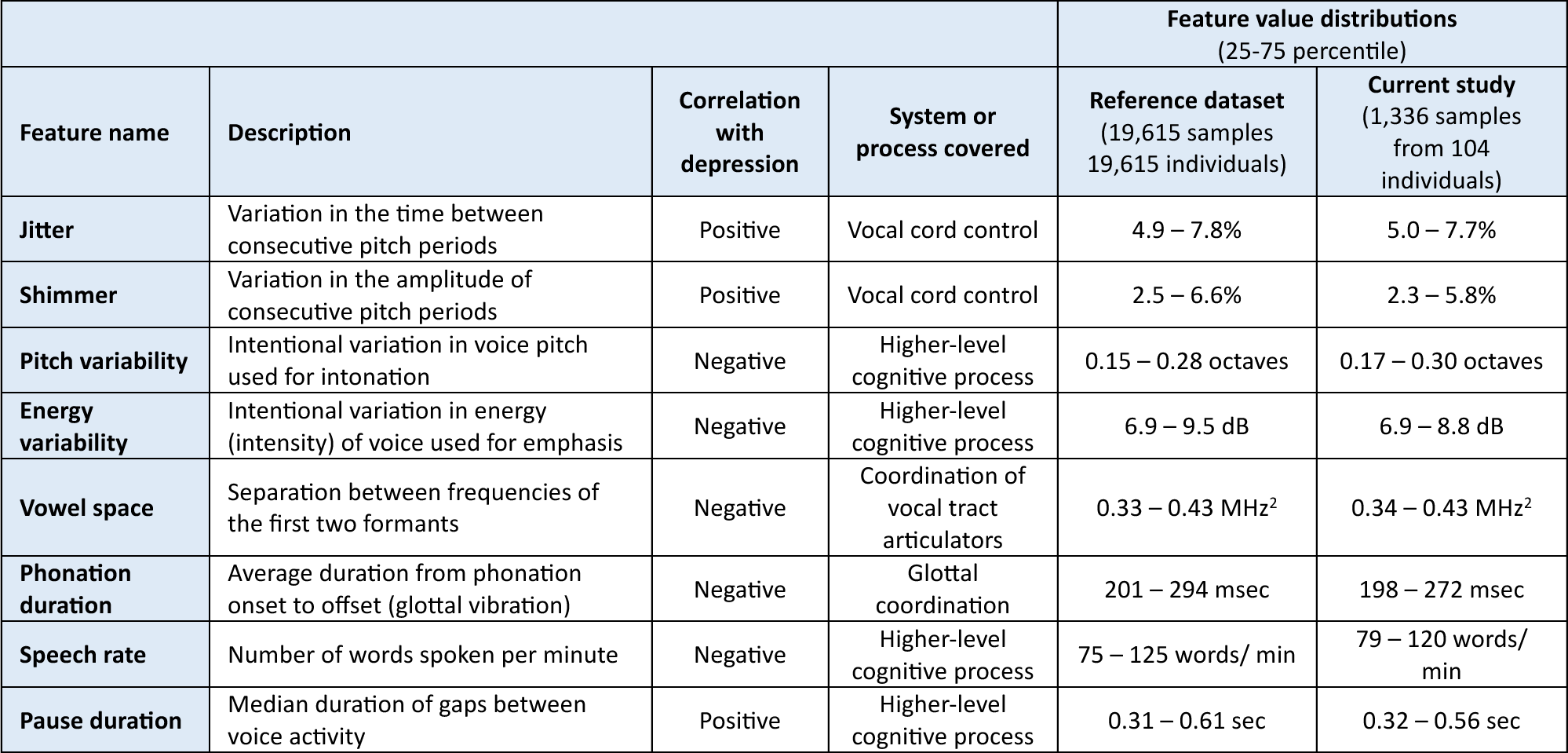
Mental health related vocal features implemented in the Mental Fitness study app. Features were selected based on available evidence from published studies on vocal biomarker research in depression. A summary score algorithm was developed using normalized values of the individual features, which were obtained from a reference dataset described in the main text. Feature value distributions obtained from the current study closely matched the reference distributions.

These features were used as inputs to the MFVB summary score using an averaging approach, resulting in an output range of 0-100 with higher scores indicating higher mental fitness (lower likelihood of elevated mental health symptoms). Before averaging, the vocal feature values were normalized using distributions (see Table 1) calculated from a large proprietary digital biobank previously acquired through clinical studies across a large number of hospital sites in India. The resulting MFVB score distribution for the India-based reference population was used to create score categories indicating relatively high, medium, or low MFVB scores. The score ranges were labeled to allow easier interpretation for the user as “Excellent”, “Good”, and “Pay Attention”.

### Study design

This study was designed and reported in accordance with STARD (57), and aimed to demonstrate that MFVB-based mental health symptom severity information could be obtained in consumer-grade products from users that are not guided or trained by a coordinator or clinician. Participants received a 1-page study instruction document that included details on how to install and set up the Mental Fitness app and how to get best results from the recordings. An online video with explanation of how to properly use the app and interpret the results was also made available. Product and study support was available to participants via email if needed, although few participants required assistance.

Participants were recruited from Cognitive Behavior Institute (Pittsburgh, PA), a provider of mental health counseling to people with a range of mental disorders, including depression, anxiety, trauma and stress, obsessive compulsive disorder, and others. The study was advertised via an email campaign that included all patients with either a recent or upcoming counseling appointment. The advertisement included a link to an online consent form where patients were able to review study details, including potential benefits and risk, incentives, and participant expectations. Patients that wished to participate could provide electronic consent and leave a contact email address, which was used by the study team to email the study instruction document described above.

The study flow is further illustrated in Fig. 1. Consent and onboarding questionnaires are hosted online through the SurveyLex platform (58). Completion of the onboarding survey was used to define Day 1 for each participant. At the end of week 4, participants received a second online questionnaire as their final study activity. Participants were informed that the study app was intended for daily use in voice journaling, although app use was self-guided.

**Figure 1.**
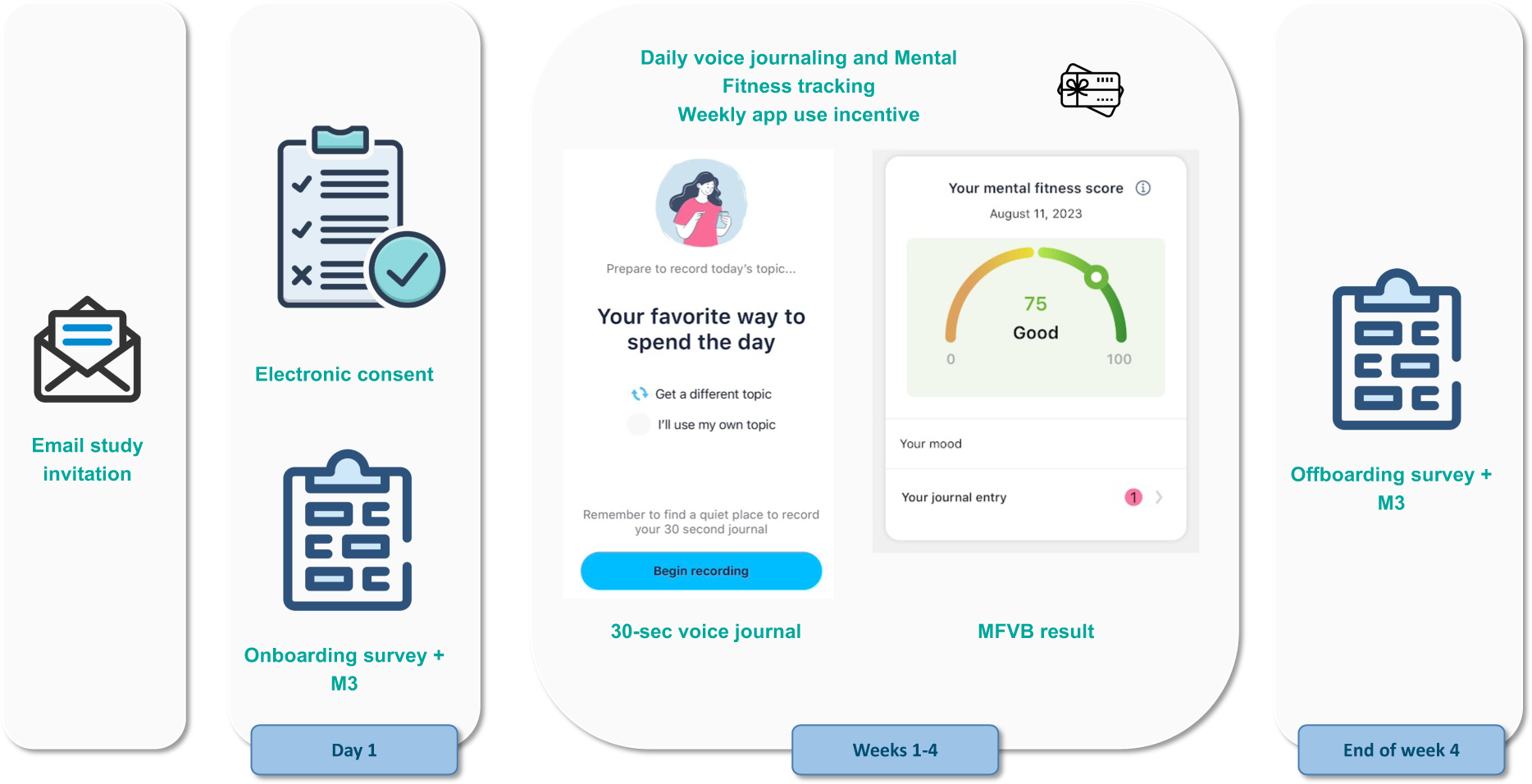
Study design and participant flow. Patients under treatment for mental health disorders at the study site receive an email invitation for the study with a link to the online consent form. After consenting and providing contact information, they receive study instructions containing a link to the onboarding survey and guidance for how to use the Mental Fitness study app. Participants use the study app for voice journaling and Mental Fitness tracking on their own for 4 weeks and receive weekly gift card incentives if they use the app at least 4 times per week. At the end of the study, they receive an offboarding survey.

Participants received a $50 Amazon gift card upon completion of the onboarding and also the final questionnaire. To incentive app use, a $15 gift card was provided on a weekly basis to participants that used the app at least 4 times during the preceding week. Participants that did not meet this requirement did not receive any reminders to promote app usage during the study. App usage was monitored by the study team using dashboards hosted on the Sonde Health infrastructure.

The 4-week per-participant duration of the study was chosen to support the study goals by allowing participants sufficient time to use the app so that they could provide meaningful feedback. It also provides a reasonable amount of time to study engagement and retention. Finally, it provides the opportunity to obtain two self-reported mental health questionnaires with sufficient separation in time, each of which can be associated with vocal biomarker results from voice recordings conducted around the time of the questionnaires.

Participants that completed both onboarding and final survey and also used the study app at least 8 times during the study were invited to participate with an extension phase of similar design but reduced incentive structure. The findings from this extension phase will be presented in future reports.

### Endpoints

MFVB scores were compared to self-reported mental health symptoms using the M3 Checklist. The M3 results include a score and symptom severity category, including normal, mild, moderate and severe; these scores and categories are provided for overall mental health and in each of the four mental health categories of depression, anxiety, post-traumatic stress disorder (PTSD), and bipolar. MFVB scores are compared to M3 severity categories. Severity categories of moderate and severe will be referred to as “elevated” vs. normal and mild as “lower”.

The primary endpoint is the Relative Risk Ratio (RR Ratio) based on relative risk (RR) estimates for elevated overall M3 symptom severity in MFVB ranges of 0-69 (“pay attention”) vs. 80-100 (“excellent”). RR is calculated as the prevalence of elevated symptom severity within a specified MFVB range divided by the analogous prevalence for the reference cohort, which will in general be the analysis cohort (defined in the Statistical section below), unless otherwise specified. An RR Ratio of 1.0 indicates that the MFVB scores have no ability to differentiate mental symptom severity risk. The primary endpoint is also computed for the four mental health domains mentioned above and also for subgroups based on demographic, clinical, and engagement-related factors^1^.

RR Ratio based on relative risk was selected as the primary endpoint instead of other measures like accuracy, sensitivity, specificity, positive/ negative predictive value as these other measures are more typically applied to diagnostic or screening instruments, which is not consistent with the intended use of the MFVB scores. Rather than providing a prediction about the likely absence or presence of mental health symptoms with a binary outcome, RR provides a more nuanced way to convey increasing or decreasing likelihood of elevated symptoms. A lower RR is not a prediction that the user does not have such symptoms, but rather that their vocal characteristics are more consistent with a reduced likelihood, and vice versa for a higher RR result. The middle MFVB score range is anticipated to display RR estimates near 1, conveying that such MFVB scores imply the users voice contains no evidence to suggest a higher or lower risk of elevated symptoms relative to the reference population.

Secondary endpoints are the engagement levels with the study app, measured as number of app sessions in study week 1, 2, 3 and 4; as well as retention in week 4, defined as the percentage of participants that use the study app at least once in week 4. Engagement groups were defined based on total app sessions: high engagement was defined as 16 or more sessions (average of 4 per week, required for receiving the weekly app use incentive), medium engagement as 8-15 sessions, low engagement as fewer than 8 sessions.

Secondary endpoints also include participant feedback as collected via the end of study questionnaire at the end of week 4, which includes multiple choice questions regarding usability and helpfulness of the app and free-text responses, which will be summarized by theme. Both engagement and feedback will be assessed in the same subgroups as mentioned in the primary endpoint description to uncover potential variations in performance or feasibility of the tool within these subgroups.

### Eligibility criteria

Patients treated at the study site were eligible for the study if they had at least one clinician-verified symptom of depression according to DSM-5 (59), 14 years or older, and have English as their first language or have conversational proficiency in English as judged from the audio recordings. Participants must own a smartphone device and willing to install and use the Mental Fitness app.

Potential participants were excluded if having a diagnosis of severe cognitive disability precluding informed consent, diagnosis of dementia, schizophrenia, or a speech disorder (e.g., apraxia, dysarthria), or use of certain medications. Although most medications are not formally evaluated for effects on voice and speech, psychiatric, metabolic or other physiologic side effects can occur (10–13). In the context of this study, a short list of psychiatric medication that may have such side effects was excluded, including first-generation antipsychotics (chlorpromazine, haloperidol, loxapine, thioridazine), two or more second-generation antipsychotics (amisulpride, aripiprazole, clozapine, iloperidone, lurasidone, olanzapine, paliperidone, quetiapine, risperidone, ziprasidone, or use of high-dose benzodiazepines (diazepine>=30 mg/day, clonazepam>=2 mg/day, lorazepam>=4 mg/day, alprazolam>=2 mg/day). Additional exclusion criteria were substance abuse, defined as any of the following behaviors in the prior 12 months: 5+ alcoholic drinks in a single day, use of prescription drugs for non-medical use, use of illicit drugs, to avoid potential confounding effects on vocal production (60,61).

Because participants were recruited via email advertisements, we relied on participant self-report in the onboarding survey to verify the exclusion criteria, in particular the criteria involving medical diagnoses and medication use. Language ability and speech disorders were judged from manual quality control on the voice recordings from the study app. Any participant who met any of the exclusion criteria was not removed from the study, but their data was omitted from the analysis cohort (described in the Statistical section below).

### Assessments and data collection

Voice recordings were collected through the Sonde Mental Fitness app on participants personal smartphone device. These were 30-second recordings of responses to predefined randomized prompts or the participant’s own topic. The voice journals were transcribed and logged in the app for the user, but transcriptions were not part of the study data set and not considered in any analysis. Acoustic features were extracted from the voice recordings and used to calculate MFVB scores and categories (see Supplemental Material). The number of voice recordings per participant varied as app use was self-directed.

SurveyLex, an online questionnaire and voice recording platform for research use (58), was used to collected various questionnaires:

- Onboarding questionnaire including demographic (gender, age, race, ethnicity), medical (excluded medications, other medication use, diagnosed medical conditions) and behavior (smoking, vaping, substance abuse, use of mental health focused apps) questions.
- M3 Checklist, collected once at onboarding and once at the end of the study in week 4.
- Feedback on the Mental Fitness app and the study. Feedback questions covered level of agreement with liking to use the study app, understanding of the results, helpfulness of the results, helpfulness as addition to treatment, and desire to keep using the study app after the end of the study. Free text responses were provided to questions about how the study app was used and how it was or was not helpful and the best and worst aspects about the app and the study.

The M3 Checklist was used to gather patient-reported mental health symptoms and includes 27 questions related to depression, anxiety, post-traumatic stress disorder (PTSD) and bipolar disorder, as well as questions related to substance abuse and impact of symptoms on work, school, social, and home life. The M3 has been validated vs. MINI mental states diagnoses and compared with existing screening instruments for mental disorders (41,42). We used the M3 due to the convenience of covering multiple mental disorders in one instrument and the ability to assess mental health symptom severity using established score ranges (normal, mild, moderate, severe) for overall transdiagnostic mental health as well for each of the 4 subdomains.

The following data was collected from patient medical records at the study site:

- Diagnosis from treating clinician, reflecting the primary focus of treatment at the time of the study. Typically, this also reflects the condition having the most significant impact on the participant’s mental health, if other conditions are present.
- Treatment duration at the study site at the time of onboarding.
- Treatment elsewhere immediately prior to treatment the study site.
- Date and results of most recent PHQ-9 administered as part of the participant’s treatment.
- Initial (start of treatment) and most recent depressive symptoms noted by the treating clinician.

Data were linked across platforms by matching patient name and email as reported in the informed consent document to medical records at the study site, survey responses, and study app account information.

### Statistical considerations

#### Analysis populations

The following cohorts are defined:

- <u>Enrolled cohort</u>: all participants that provided informed consent.
- <u>Onboarded cohort</u>: subset of the enrolled cohort which completed the onboarding questionnaire at the start of the study.
- <u>Analysis cohort</u>: subset of the onboarded cohort which completed at least 1 Mental Fitness app voice recording session within a period starting 2 weeks before onboarding and ending 2 weeks after the final study survey in week 4 and did not meet any of the exclusion criteria. This cohort allows comparison between MFVB results and mental symptoms per the study’s primary endpoint as well as the engagement analysis in the secondary endpoints.
- <u>Completer cohort</u>: subset of the analysis cohort which also completed the final study survey in week 4. This cohort allows assessment of the feedback responses per the study’s secondary endpoints.

The 2-week time window mentioned in the definition of the analysis cohort was used because some participants used the study app before completing the onboarding questionnaire (or after the offboarding questionnaire). As described below, a 2-week window is used to associate study app voice recordings to M3 surveys.

#### Statistical analyses

The association between MFVB scores and M3 results was conducted in two ways, illustrated in Fig. 2:

1. <u>Closest-MFVB</u>: The single MFVB result that was nearest in time (before or after) to the completion of the M3, but not more than 2 weeks before or after.
2. <u>Time-weighted MFVB</u>: Using all MFVB scores that were available from 2 weeks before to 2 weeks after the M3 was completed. Each MFVB result was weighted as function of its temporal proximity to the M3 with a linear decreasing function of absolute time separation (weighting of 1 for no separation, weighting of 0 for 2 weeks separation). After the initial weights were thus assigned, they were all scaled by a constant so that the weights would sum to 1.

**Figure 2.**
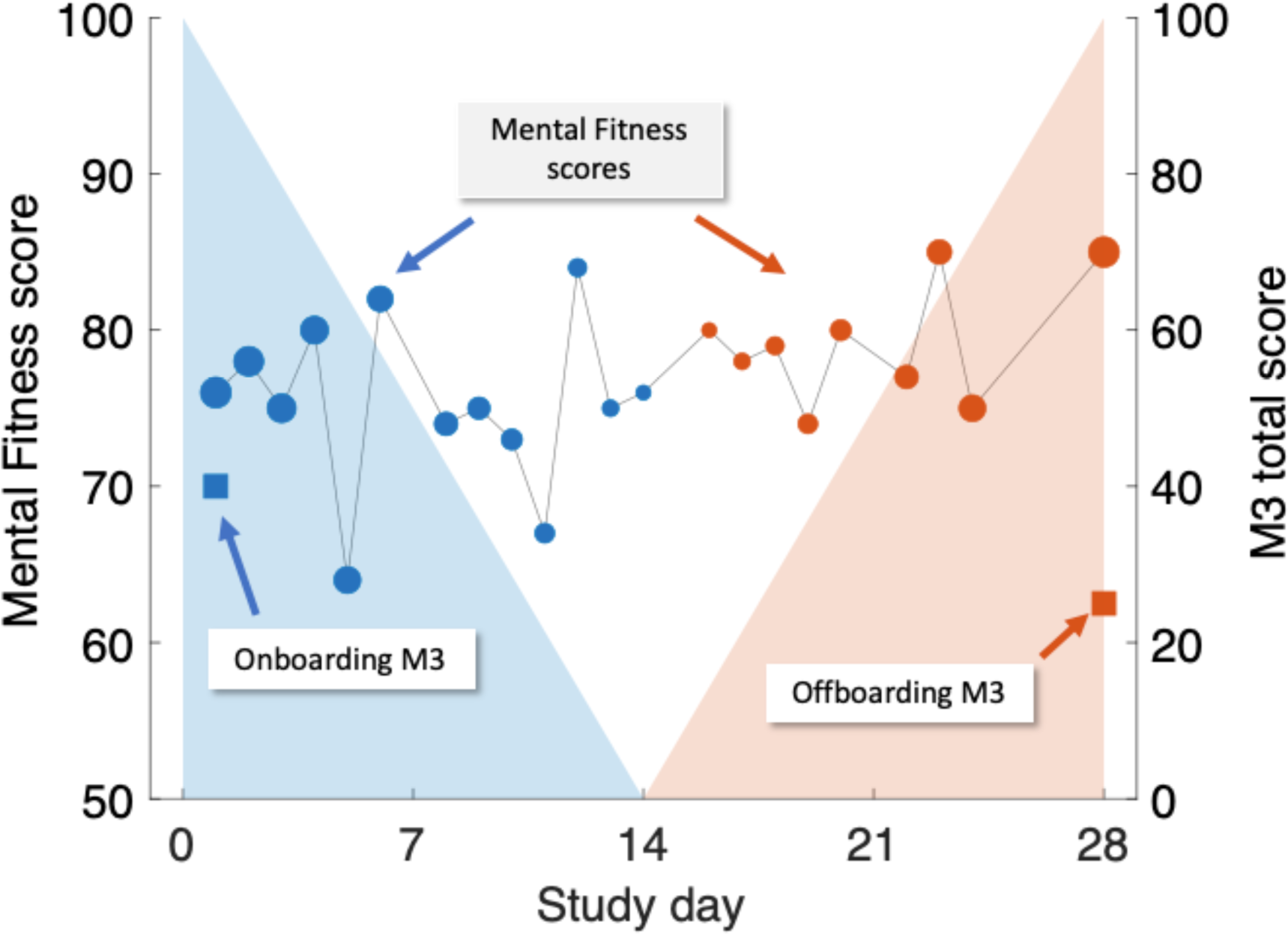
Illustration of time-weighted method to link M3 assessments (right-hand scale, square symbols) to Mental Fitness Vocal Biomarker (MFVB) scores (left-hand scale, round symbols). M3 assessments are obtained on Day 1 (blue) and Day 28 (red) and are each paired with weighted averages of MFVB scores obtained in a period from 2 weeks prior to 2 weeks after the M3 timepoint. Weighting emphasizes MFVB scores closer to the M3 time point, indicated by the size of the MFVB symbol and the decreasing shading applied around each M3 time point. Weights are normalized to sum to 1. Note that in this example, the M3 total score declines over time, paired with a positive trending MFVB score, as intended (recreation of actual participant data).

The rationale and interpretation for the closest-MFVB approach is that this demonstrates the association between a single voice recording analysis to the M3 result, which is useful for one-time or screening-like approaches.

The time-weighted approach evaluates if the association between MFVB scores and M3 results can be enhanced through repeated app usage over time. This is particularly relevant for scenarios like fitness tracking, where users engage with the application repeatedly. The chosen 2-week window aligns with the M3 survey’s duration, mirroring similar time frames in mental health questionnaires like PHQ-9 and GAD-7. This window encourages participants to consider symptoms over time, reducing the risk of momentary biases during assessments. Aggregating vocal characteristics over a 2-week period enhances alignment with survey results, offering a more comprehensive evaluation compared to a single time point measure.

The implementation of decaying weights in the time-weighted MFVB approach introduces a recency bias, assuming that M3 responses are more influenced by recent feelings and moods during the 2-week window. The linear weighting function was selected for its simplicity in incorporating this recency bias into the MFVB aggregation. Associating MFVB scores from both before and after the M3 questionnaire is suitable under the assumption that symptoms change gradually, allowing mental states after the M3 completion to remain correlated with the result for some duration. This ensures that vocal characteristics during this time window remain pertinent to the mental health assessment.

### Ethical considerations

The study was conducted in accordance with the principles of Good Clinical Practice as described by the study designs and controls previously. Furthermore, the research protocol was reviewed and approved by WCG IRB (Protocol #20220961) prior to study initiation, and the Hamilton Integrated Research Ethics Board (Project #14494) for the pilot phase at St Joseph’s Healthcare Hamilton. All participants provided electronic informed consent in the online SurveyLex platform before participating in the study. The study app and databases were hosted on Amazon Web Services cloud servers with various security mechanisms in place, described in more detail in the Supplementary Material. Participants were made aware in the informed consent information about financial incentives in the form of electronic gift vouchers upon completion of onboarding and final questionnaire ($50 in each case) and $15 for any of the 4 study weeks if they used the study app at least 4 times during that week. Total potential incentive amount was $160. Incentives were provided solely based on the completion of these study activities and were not contingent on specific outcomes.

## RESULTS

### Enrollment

The participant cohorts are illustrated in Fig. 3: a total of 147 participants from the study site provided electronic informed consent on the web application between May and August 2023. All these participants were emailed study instructions and a link to the onboarding questionnaire, which was completed by 120 participants. Of these, 115 participants met eligibility criteria, and of those 104 participants used the study app at least once during the study. This set of 104 participants forms the analysis cohort and will be used to report results, unless otherwise indicated. The offboarding survey at the end of week 4 was completed by 81 participants, forming the completer cohort.

**Figure 3.**
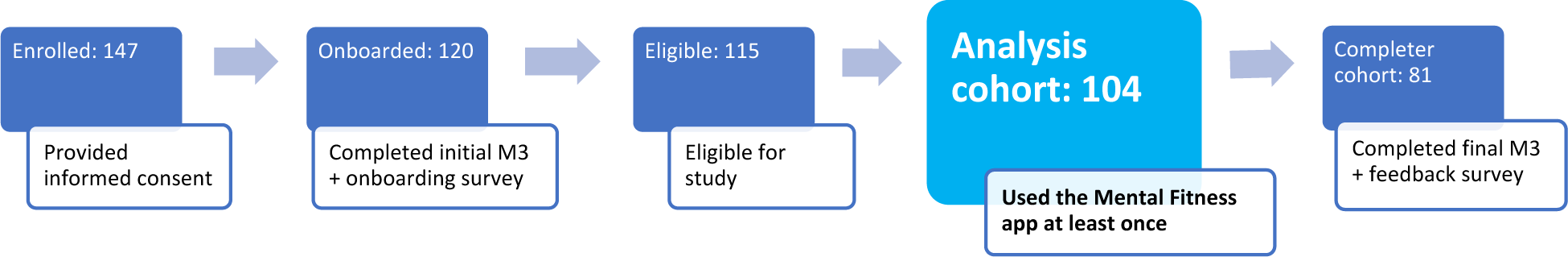
Cohort definitions and participant counts.

### Demographics and health characteristics

Demographic and health characteristics of the analysis cohort are provided in Table 2. The analysis cohort was majority female (73%); skewed to younger adults with proportion in age ranges below 30, 30-39 and over 40 of 41%, 27% and 32% (age range 16-80); mostly white (93%) and non-Hispanic (99%). Most participants have been in longer-term treatment (66% for >1 or 2 years) and were not treated elsewhere prior to their current therapy (75%). Most participants (73%) reported using prescription medications for their mental health treatment (main reported medications being fluoxetine, sertraline, escitalopram, bupropion, lamotrigine, buspirone, lisdexamfetamine, methylphenidate, venlafaxine, and duloxetine). About half of the participants (47%) reported one or more diagnosed health conditions, including obesity (19%), asthma (15%), hypertension (12%), diabetes (3%) and other conditions (19%).

**Table 2.**
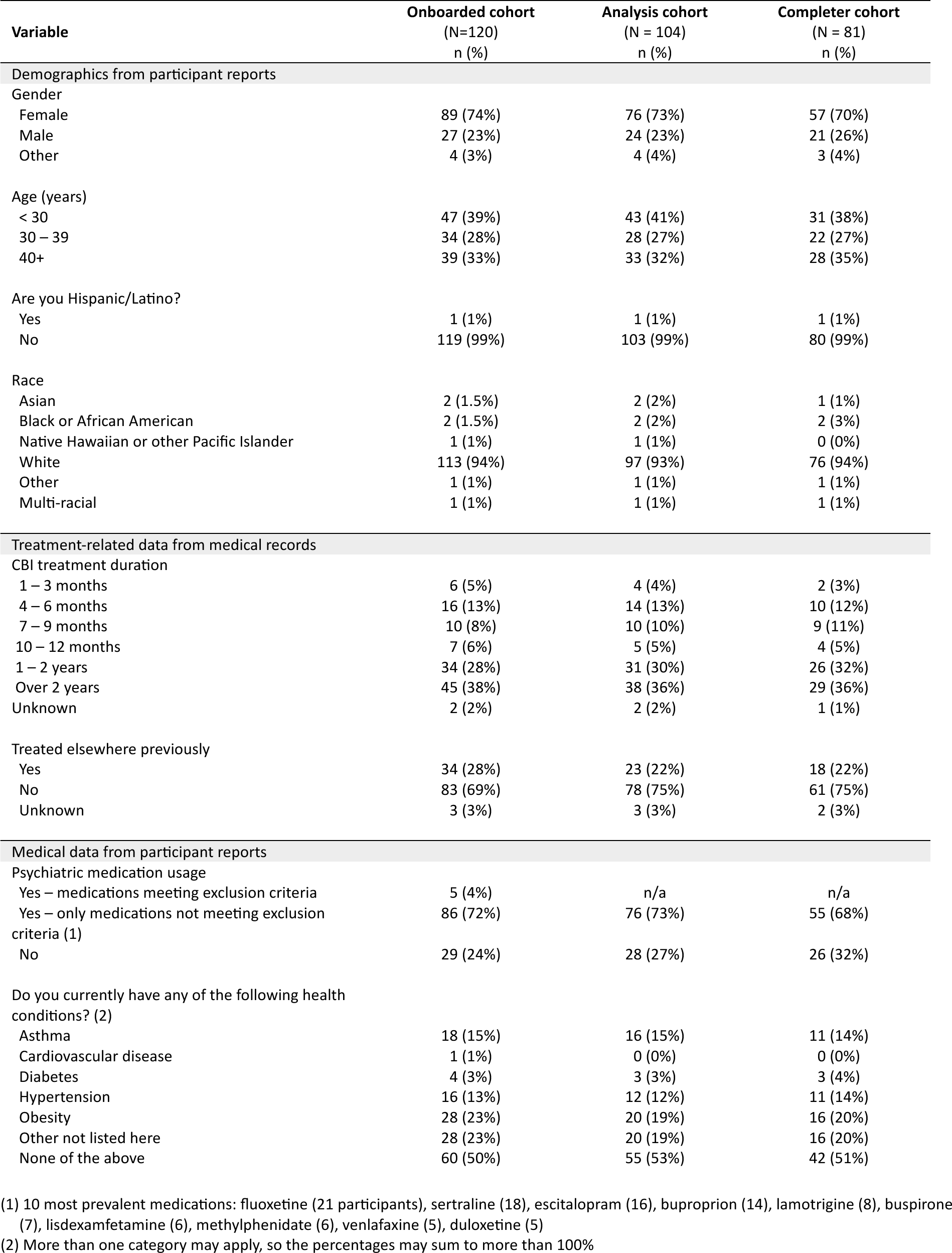
Demographics, treatment, and medical characteristics of the study participants. Information was reported by participants, except treatment information which was obtained from medical records.

| Variable | Onboarded cohort<br>(N=120)<br>n (%) | Analysis cohort<br>(N = 104)<br>n (%) | Completer cohort<br>(N = 81)<br>n (%) |
| --- | --- | --- | --- |
| Demographics from participant reports |  |  |  |
| Gender |  |  |  |
| Female | 89 (74%) | 76 (73%) | 57 (70%) |
| Male | 27 (23%) | 24 (23%) | 21 (26%) |
| Other | 4 (3%) | 4 (4%) | 3 (4%) |
| Age (years) |  |  |  |
| < 30 | 47 (39%) | 43 (41%) | 31 (38%) |
| 30 – 39 | 34 (28%) | 28 (27%) | 22 (27%) |
| 40+ | 39 (33%) | 33 (32%) | 28 (35%) |
| Are you Hispanic/Latino? |  |  |  |
| Yes | 1 (1%) | 1 (1%) | 1 (1%) |
| No | 119 (99%) | 103 (99%) | 80 (99%) |
| Race |  |  |  |
| Asian | 2 (1.5%) | 2 (2%) | 1 (1%) |
| Black or African American | 2 (1.5%) | 2 (2%) | 2 (3%) |
| Native Hawaiian or other Pacific Islander | 1 (1%) | 1 (1%) | 0 (0%) |
| White | 113 (94%) | 97 (93%) | 76 (94%) |
| Other | 1 (1%) | 1 (1%) | 1 (1%) |
| Multi-racial | 1 (1%) | 1 (1%) | 1 (1%) |
| Treatment-related data from medical records |  |  |  |
| CBI treatment duration |  |  |  |
| 1 – 3 months | 6 (5%) | 4 (4%) | 2 (3%) |
| 4 – 6 months | 16 (13%) | 14 (13%) | 10 (12%) |
| 7 – 9 months | 10 (8%) | 10 (10%) | 9 (11%) |
| 10 – 12 months | 7 (6%) | 5 (5%) | 4 (5%) |
| 1 – 2 years | 34 (28%) | 31 (30%) | 26 (32%) |
| Over 2 years | 45 (38%) | 38 (36%) | 29 (36%) |
| Unknown | 2 (2%) | 2 (2%) | 1 (1%) |
| Treated elsewhere previously |  |  |  |
| Yes | 34 (28%) | 23 (22%) | 18 (22%) |
| No | 83 (69%) | 78 (75%) | 61 (75%) |
| Unknown | 3 (3%) | 3 (3%) | 2 (3%) |
| Medical data from participant reports |  |  |  |
| Psychiatric medication usage |  |  |  |
| Yes – medications meeting exclusion criteria | 5 (4%) | n/a | n/a |
| Yes – only medications not meeting exclusion criteria (1) | 86 (72%) | 76 (73%) | 55 (68%) |
| No | 29 (24%) | 28 (27%) | 26 (32%) |
| Do you currently have any of the following health conditions? (2) |  |  |  |
| Asthma | 18 (15%) | 16 (15%) | 11 (14%) |
| Cardiovascular disease | 1 (1%) | 0 (0%) | 0 (0%) |
| Diabetes | 4 (3%) | 3 (3%) | 3 (4%) |
| Hypertension | 16 (13%) | 12 (12%) | 11 (14%) |
| Obesity | 28 (23%) | 20 (19%) | 16 (20%) |
| Other not listed here | 28 (23%) | 20 (19%) | 16 (20%) |
| None of the above | 60 (50%) | 55 (53%) | 42 (51%) |

(1) 10 most prevalent medications: fluoxetine (21 participants), sertraline (18), escitalopram (16), bupropion (14), lamotrigine (8), buspirone (7), lisdexamfetamine (6), methylphenidate (6), venlafaxine (5), duloxetine (5)
(2) More than one category may apply, so the percentages may sum to more than 100%

### Clinical presentation

Psychiatric diagnoses were extracted from participant medical records at the study site and grouped into high-level categories as indicated in Table 3. Anxiety-related diagnoses were the most common (38% of the analysis cohort), followed by trauma and stress-related disorders (31%), and depression disorders (20%). The most prevalent specific diagnoses were generalized anxiety disorder (22% of the analysis cohort), major depressive disorder (16%), adjustment disorder (19%), obsessive-compulsive disorder (11%), and post-traumatic stress disorder (PTSD, 10%). Various other conditions were also present in smaller numbers, e.g., attention-deficit hyperactivity disorder, bipolar disorder, and borderline personality disorder.

**Table 3.** Psychiatric diagnoses obtained from patient medical records. These reflect the disorder which most impact participant functioning, for individuals with multiple diagnosed disorders.

|  | Consented<br>N = 147 | Onboarded<br>N = 120 | Analysis<br>N = 104 | Completer<br>N = 81 |
| --- | --- | --- | --- | --- |
| **Anxiety** |  |  |  |  |
| Generalized anxiety disorder | 37 | 26 | 23 | 18 |
| Obsessive-compulsive disorder | 11 | 11 | 11 | 9 |
| Social anxiety | 3 | 3 | 3 | 3 |
| Anxiety – other | 3 | 2 | 2 | 0 |
| Panic disorder | 1 | 1 | 1 | 1 |
| **All anxiety disorders** | **55** | **43** | **40** | **31** |
| **Depression** |  |  |  |  |
| Major depressive disorder | 25 | 21 | 17 | 13 |
| Depression - other | 5 | 4 | 2 | 2 |
| Persistent depressive disorder | 2 | 2 | 2 | 1 |
| **All depressive disorders** | **32** | **27** | **21** | **16** |
| **Trauma- and stress-related disorders** |  |  |  |  |
| Adjustment disorder | 23 | 21 | 20 | 17 |
| Post-traumatic stress disorder | 13 | 12 | 10 | 9 |
| Trauma – other | 3 | 2 | 2 | 2 |
| **All trauma- and stress-related disorders** | **39** | **35** | **32** | **28** |
| **Other** |  |  |  |  |
| Attention-deficit hyperactivity disorder | 8 | 4 | 4 | 3 |
| Bipolar disorder | 4 | 3 | 2 | 1 |
| Borderline personality disorder | 3 | 3 | 2 | 1 |
| Feeding or eating disorder | 1 | 1 | 0 | 0 |
| Autism | 1 | 1 | 0 | 0 |
| Unspecified | 4 | 3 | 3 | 1 |
| **All other disorders** | **21** | **15** | **11** | **6** |

Among consented participants that failed to onboard onto the study, a relatively large number included participants with diagnoses of generalized anxiety disorder (11, or 30% of the consented group with this diagnosis) and attention-deficit hyperactivity disorder (4, or 50%). Onboarded participants that never used the study app or were not eligible and therefore not included in the analysis cohort were not concentrated in any particular diagnosis, being somewhat high only in major depressive disorder (4, or 19% of the onboarded group with this diagnosis). Most participants (80%) also completed the offboarding questionnaire (completer cohort) and no major differences between diagnostic categories in this regard were noted.

Table 4 offers insights into mental health symptom severity through M3 Checklists, encompassing 104 participants and 185 M3 assessments (5 M3 assessments could not be linked to any participant and were excluded from analysis). Results are presented at onboarding for both the analysis and completer cohorts, allowing for direct comparisons with offboarding outcomes available exclusively from the completer cohort. At onboarding, the analysis cohort presented the following severity distribution: normal (0%), mild (31%), moderate (48%), and severe (21%). The cumulative prevalence of elevated (moderate-to-severe) symptoms was 69%. Examining mental health categories, elevated symptoms were prevalent in 53% for depression, 30% for anxiety, 27% for PTSD, and 11% for bipolar.

**Table 4.**
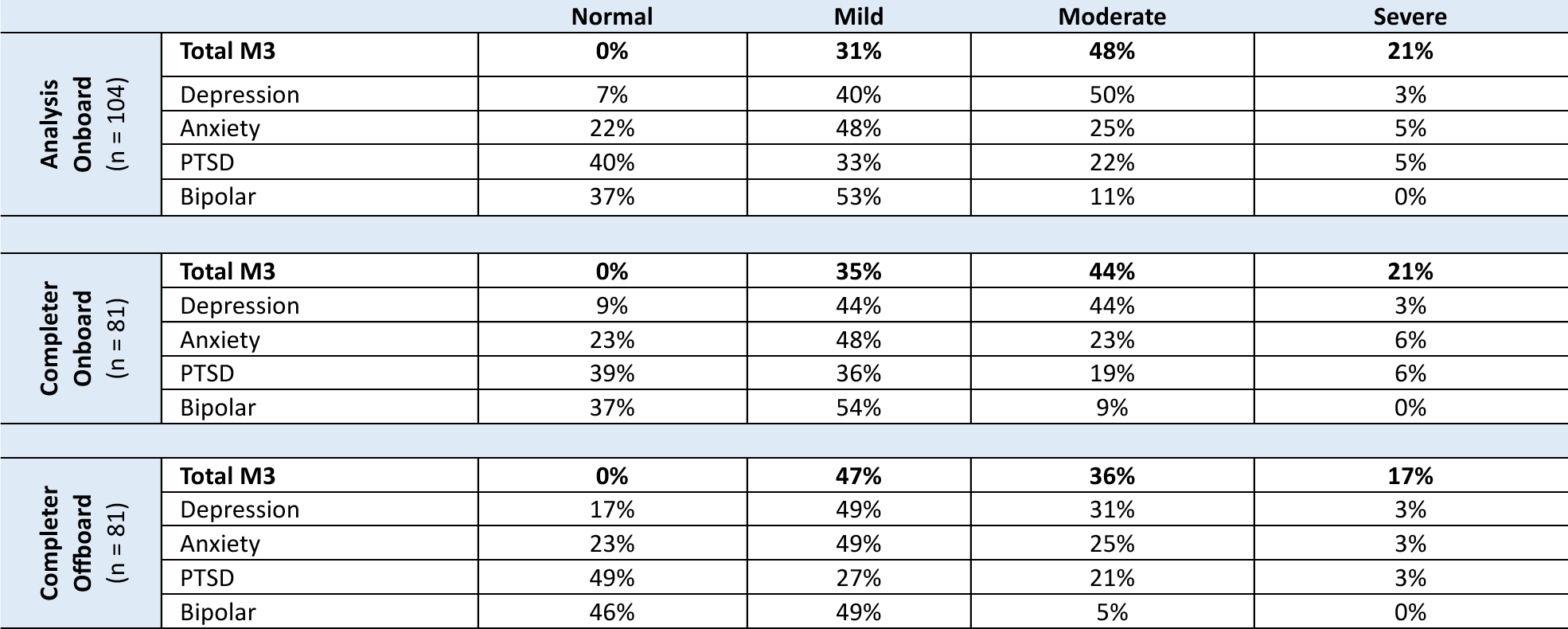
Mental health symptom severity prevalence reported using the M3 Checklist at onboarding (analysis and completer cohorts) and offboarding. Comparison of symptom categories between onboard and offboard should refer to the completer cohort, as 21 out of 104 participants in the analysis cohort did not complete the offboarding survey. These non-completers had somewhat higher symptom severity vs. the completer cohort (see Main text).

In approximately one-third of participants, elevated symptoms were observed across multiple mental health categories, indicating a high overall symptom burden (100% of these with elevated total symptom severity). The most prevalent comorbid symptom categories involved depression, anxiety, and PTSD, with additional occurrences of elevated bipolar symptoms in certain cases. Depression with anxiety and depression with PTSD was also observed. Notably, elevated depressive symptoms were widespread, while elevated bipolar symptoms were relatively uncommon, except for a small subset exhibiting elevated symptoms across all categories. Consequently, the presence of elevated bipolar symptoms emerged as the primary predictor for elevated overall mental symptom burden and exhibited the highest average number of comorbid symptom categories. These findings underscore the importance of comprehensive mental health assessment, enabled by transdiagnostic tools like the M3.

Symptom severity at onboarding was correlated with the likelihood to complete the offboarding survey: completers vs. non-completer (participants from the analysis cohort that are not included in the completer cohort) demonstrated the following elevated symptom severity distribution: Total (65% vs. 81%), depression (47% vs. 71%), anxiety (29% vs. 33%), PTSD (27% vs. 29%), and bipolar (9% vs. 14%).

Symptom severity reported in the offboarding survey at the end of week 4 indicates a notable reduction in elevated symptom severity within the completer cohort over the study period. The prevalence decreased from 65% at onboarding to 53% at offboarding, reflecting a 13% reduction—suggestive of the potential impact of treatment provided at the study site. The average reduction in elevated symptom severities across the four subcategories was 5%, with the most substantial improvement observed in depression (13%), while the lowest reductions were noted in PTSD (2%) and anxiety (1%). These improvements were also noted on functional impairment outcomes of the M3: 25-30% of participants reported improvements in functioning at work or school and relationships with friends or family. These numbers rose to 35-40% for those participants that reported significant impairment in functioning on their initial M3.

### Voice sample recordings

Participants conducted 1,336 app sessions with voice recordings during their 4-week study period, resulting in an average 12.8 sessions per participant, or 3.2 per week – nearly exactly once every other day. Additional detail regarding app use will be provided in the Participant Engagement section below. Vocal feature distributions from the study data set were compared with those from the reference dataset that was used to develop the MFVB scoring algorithm and were found to be in close alignment (see Table 1).

### Relationship of vocal biomarker to mental health symptom severity

Out of 185 M3 assessments, 177 were included in analysis of the primary endpoint due to having 1 more associated MFVB results within the 2-week time window. The prevalence or risk of elevated mental health symptom severity across this dataset was 62% (110 out of 177), defining the Relative Risk (RR) level of 1.00.

Visual representation of the alignment between MFVB score categories and total symptom scores from the M3 assessment is shown in Fig. 4, using the closest-MFVB and time-weighted MFVB approach. Both approaches indicate that M3 scores and thus symptom severity distributions increase from MFVB categories of “Excellent” to “Good” to “Pay Attention”, as intended.

**Figure 4.**
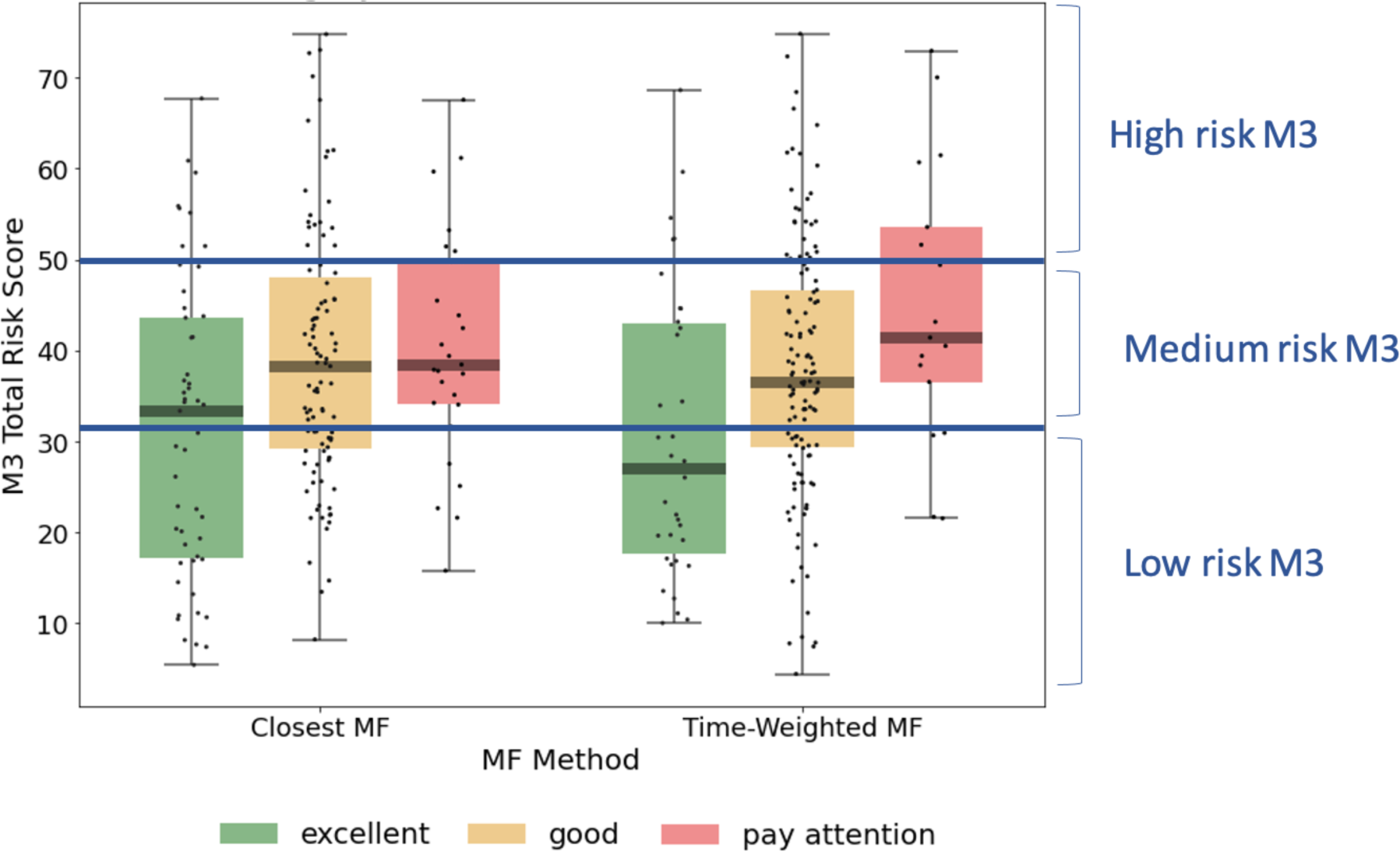
M3 total risk score distributions by MFVB score category, indicating the correlation between vocal analysis results and participant reported mental health symptom severity. The left-hand side of the panel associated the MFVB result closest in time to the M3, whereas the right-hand side of the panel uses a time-weighted average of MFVB results within 2 weeks of the M3 (see Methods). Results in Table 5 are calculated on the same data as displayed here.

**Table 5.** Relative Risk (RR) for MFVB score categories compared to the analysis cohort (“all”) for total mental health symptoms and symptom categories of depression, anxiety, PTSD and bipolar. Results are shown for the association of a single MFVB score to the closest M3 assessment (closest-MFVB) and for the association of potentially multiple MFVB scores within a 2-week window around the M3 assessment (time-weighted MFVB). See main text and Fig. 2 for details on the calculation methodology. Elevated M3: moderate or severe; lower M3: normal or mild. RR Ratio is the ratio of the RR estimate for Pay Attention vs. Excellent categories.

|  |  | Closest-MFVB |  |  |  | Time-weighted MFVB |  |  |  |
| --- | --- | --- | --- | --- | --- | --- | --- | --- | --- |
|  | MFVB score category | Elevated M3 count | Lower M3 count | RR (95% C.I.) | RR Ratio (95% C.I.) | Elevated M3 count | Lower M3 count | RR (95% C.I.) | RR Ratio (95% C.I.) |
| M3 Total | Excellent | 26 | 25 | 0.82 (0.61, 1.10) | 1.53 (1.09-2.14)<br>$p=0.0138$ | 13 | 21 | 0.62 (0.40, 0.96) | 2.00 (1.21-3.30)<br>$p=0.0068$ |
|  | Good | 63 | 36 | 1.02 (0.85, 1.24) |  | 84 | 42 | 1.07 (0.91, 1.27) |  |
|  | Pay attention | 21 | 6 | 1.25 (0.99, 1.58) |  | 13 | 4 | 1.23 (0.92, 1.64) |  |
|  | All | 110 | 67 | 1.00 |  | 110 | 67 | 1.00 |  |
| Depression | Excellent | 17 | 34 | 0.73 (0.48, 1.11) | 1.78 (1.08-2.93)<br>$p=0.0237$ | 10 | 24 | 0.64 (0.37, 1.11) | 2.60 (1.45-4.66)<br>$p=0.0013$ |
|  | Good | 48 | 51 | 1.06 (0.82, 1.37) |  | 58 | 68 | 1.01 (0.79, 1.29) |  |
|  | Pay attention | 16 | 11 | 1.29 (0.91, 1.84) |  | 13 | 4 | 1.67 (1.23, 2.28) |  |
|  | All | 81 | 96 | 1.00 |  | 81 | 96 | 1.00 |  |
| Anxiety | Excellent | 9 | 42 | 0.68 (0.36, 1.29) | 1.47 (0.62-3.51)<br>$p=0.3865$ | 7 | 27 | 0.79 (0.39, 1.60) | 2.00 (0.84-4.78)<br>$p=0.1188$ |
|  | Good | 30 | 69 | 1.17 (0.79, 1.72) |  | 32 | 94 | 0.98 (0.66, 1.44) |  |
|  | Pay attention | 7 | 20 | 1.00 (0.50, 1.98) |  | 7 | 10 | 1.58 (0.85, 2.95) |  |
|  | All | 46 | 131 | 1.00 |  | 46 | 131 | 1.00 |  |
| PTSD | Excellent | 11 | 40 | 0.87 (0.48, 1.55) | 1.72 (0.84-3.52)<br>$p=0.1401$ | 5 | 29 | 0.59 (0.25, 1.38) | 2.40 (0.85-6.75)<br>$p=0.0971$ |
|  | Good | 23 | 76 | 0.93 (0.60, 1.45) |  | 33 | 93 | 1.05 (0.71, 1.55) |  |
|  | Pay attention | 10 | 17 | 1.49 (0.86, 2.59) |  | 6 | 11 | 1.42 (0.71, 2.84) |  |
|  | All | 44 | 133 | 1.00 |  | 44 | 133 | 1.00 |  |
| Bipolar | Excellent | 3 | 48 | 0.74 (0.22, 2.49) | 1.26 (0.22-7.08)<br>$p=0.7937$ | 2 | 32 | 0.74 (0.18, 3.12) | 0.39 (0.02-7.68)<br>$p=0.5348$ |
|  | Good | 9 | 90 | 1.15 (0.52, 2.56) |  | 12 | 114 | 1.20 (0.58, 2.51) |  |
|  | Pay attention | 2 | 25 | 0.94 (0.23, 3.89) |  | 0 | 17 | 0.34 (0.02, 5.48) |  |
|  | All | 14 | 163 | 1.00 |  | 14 | 163 | 1.00 |  |

RR was determined within MFVB score categories of Excellent, Good, and Pay Attention. Table 5 outlines the RR estimates and 95% confidence intervals (omitted here for brevity) for these MFVB score categories as 0.82, 1.02, and 1.25 when using the closest-MFVB method and 0.62, 1.07, and 1.23 when using the time-weighted method.

Notably, both methods show that the “Excellent” and “Pay Attention” MFVB categories reflect RR below and above 1.00 respectively, as originally intended, as well as an RR near 1 for the MFVB score category of “Good”. The RR Ratios between Pay Attention and Excellent MFVB score categories for these two methods are statistically significant at 1.53 (1.09-2.14, p=0.0138) and 2.00 (1.21-3.30, p=0.0068), respectively, meeting the primary endpoint of the study. A principal effect of the time-weighted versus the closest-MFVB method is an increased distinction in RR away from 1, presumably benefiting from “multiple looks” at the MFVB results across the 2-week aggregation period.

The same principles observed for the total M3 can be extended to the subcategories for depression, anxiety, PTSD, and bipolar (Table 5). MFVB efficacy for depression is more pronounced compared to total symptoms: this heightened distinction may be attributed to the selection of vocal features, which were primarily chosen for their established association with depression. RR Ratios for closest and time-weighted methods are both statistically significant at 1.78 (1.08-2.93, p=0.0237) and 2.60 (1.45-4.66, p=0.0013), respectively. Results for anxiety and PTSD indicate consistent RR trends despite wider confidence intervals and non-significant RR Ratio due to a lower prevalence of elevated symptoms in these categories. It is particularly challenging to accurately assess RR for bipolar due to the limited total number of reported cases of elevated symptom severity (only 14 M3 results fall into this category).

Additionally, hypomanic patients may manifest a mixture of vocal qualities, some of which are normally associated with strong mental health.

### Participant engagement with the Mental Fitness study app

As indicated in Table 6, participants engaged with the study app an average of 12.8 total times over the course of the 4-week study period, with 70% of users remaining active in week 4. These average usage patterns can be further examined by categorizing engagement into three levels, as described in the Methods section.

**Table 6.** Participant engagement during the 4-week study period, measured as average weekly sessions and retention in week 4. Engagement varies considerably between participants and is summarized in groups with high (16+ sessions total), medium (8-15 sessions total), and low (1-7 sessions total) engagement.

| Engagement group | Count (%) | Average sessions per week |  |  |  | Week 4 retention |
| --- | --- | --- | --- | --- | --- | --- |
|  |  | Week 1 | Week 2 | Week 3 | Week 4 |  |
| All | 104 (100) | 3.9 | 3.4 | 3.0 | 2.5 | 70% |
| High | 40 (38) | 5.7 | 5.5 | 4.9 | 5.0 | 100% |
| Medium | 34 (33) | 3.7 | 3.3 | 3.0 | 1.6 | 71% |
| Low | 30 (29) | 1.7 | 0.9 | 0.6 | 0.3 | 14% |

The high engagement category included 38% of participants, with a consistent 5-6 app sessions each week, 100% retention in week 4 and an average of 21.1 sessions in total (approximately 10 minutes of voice recordings over the study period). The medium engagement group (8-15 sessions) comprised 33% of participants. This group typically utilized the app 3-4 times per week with a gradual decrease over time, had 71% retention rate in week 4, and an average of 11.6 sessions overall. The remaining 29% of participants exhibited low engagement and had minimal app usage, even in the initial study week; only 14% of these users were retained in week 4, with an average of 3.5 total app uses. The engagement groups thus differed both on total app usage but also persistence of use over time.

To find potential reasons for these very different levels of engagement, cohorts defined by the engagement groups were constructed and assessed for variations in demographics, health characteristics, and clinical presentation. Statistical significance was assessed in each case using chi-square test and 5% significance level. The only statistically significant factor was age (p=0.0131), with older age groups displaying higher levels of engagement. The oldest age group (40+ years) comprised 45% of the high engagement group, 26% of the medium engagement group, and 20% of the low engagement. While the middle age group (30-39 years) trended similarly, for the youngest age group (<30 years) the analogous proportions were 20%, 53%, and 57%, revealing an opposite trend.

Other factors, including male gender (p=0.1273), absence of prescription medication use in mental health treatment (p=0.3157), lower symptom severity (p=0.4425, comparing completer and non-completer participants), and having a psychiatric diagnosis other than anxiety-related, exhibited positive trends toward increased engagement, although statistical significance was not reached. Notably, age and these non-significant factors, which trended positively with higher engagement, were also linked to lower symptom severity. This suggests that symptom severity may be a primary determinant of engagement, with higher symptom levels associated with reduced engagement and vice versa. This aligns with the expectation that individuals experiencing more severe symptoms might be less engaged.

### Participant feedback

Participant agreement with five statements presented in the offboarding questionnaire at the conclusion of week 4 is summarized in Table 7. This questionnaire was completed by 80 participants, 77% of the analysis cohort. The high response rate reduces potential responder bias in the response analysis.

**Table 7.** Participant level of agreement with statements in the feedback portion of the offboarding survey; 80 participants included (completer cohort, except 1 participant that did not provide feedback responses).

|  | I like being able to check my Mental Fitness using a voice recording on my smartphone. | I understand what the mental fitness scores calculated from my voice mean. | The mental fitness scores are helpful to me in understanding how I'm doing. | Using the mental fitness app has been a helpful addition to my treatment at CBI. | I intend to (or would like to) keep using the mental fitness app to track how I'm doing in the future. |
| --- | --- | --- | --- | --- | --- |
| (5) Completely agree | 34 | 23 | 14 | 20 | 28 |
| (4) Somewhat agree | 33 | 38 | 42 | 26 | 32 |
| (3) Neither agree nor disagree | 8 | 5 | 9 | 28 | 10 |
| (2) Somewhat disagree | 5 | 11 | 13 | 3 | 7 |
| (1) Completely disagree | 0 | 3 | 2 | 3 | 3 |
| *Proportion somewhat or completely agree* |  |  |  |  |  |
| **All participants** | **81%** | **73%** | **67%** | **55%** | **72%** |
| High engagement | 92% | 82% | 79% | 71% | 87% |
| Med engagement | 75% | 64% | 54% | 50% | 64% |
| Low engagement | 65% | 71% | 65% | 29% | 53% |
| **Average response score (1-5 range)** | 4.1 | 3.7 | 3.5 | 3.6 | 3.8 |

In general, a substantial majority of participants expressed partial or full agreement with all statements. The highest level of agreement (81%) was in response to the statement expressing contentment with the capability to assess Mental Fitness through voice recordings on their smartphone, underscoring the favorable reception of this activity and application among participants. The lowest level of agreement (55%) was noted in response to the statement asserting that the Mental Fitness app constituted a beneficial adjunct to their treatment at CBI. Here, a relatively large proportion of participants indicated a neutral opinion. It is noteworthy that neither the design of the Mental Fitness app nor the study itself inherently positioned it as a supplement to mental health treatment. Consequently, the observed level of agreement can in fact be considered rather high. Evidently, about half of the study participants organically discovered meaningful ways to incorporate the app into their treatment regimens. The remaining three statements (pertaining to comprehension of MFVB scores, the perceived utility of MFVB scores for self-assessment, and the intention to continue using the Mental Fitness app in the future) received partial or full agreement from approximately 70% of participants.

Participant responses to the statements were also analyzed based on level of engagement to determine if satisfaction correlated with usage (Table 7). Results showed that participants in the high engagement group agreed more frequently, with 82% somewhat or completely agreeing on average across all statements. Using the same calculation, the medium engagement group had a 61% agreement rate, while the low engagement group had a 57% rate. Interestingly, the statement about understanding the MFVB scores showed a modest 9% difference between the high and low engagement groups, indicating that level of understanding of the scores wasn’t a significant factor driving app usage. Conversely, the statement about the app being a helpful addition to treatment at CBI had the most substantial differences between engagement groups, ranging from 71% agreement in the high engagement group to 50% in the medium engagement group and 29% in the low engagement group. This suggests that the perceived value of the app as an addition to treatment was closely tied to consistent app usage. Overall, engagement and satisfaction were strongly correlated.

### Free response feedback

The offboarding survey at the end of week 4 also included several free response questions where participants could provide more detail about how they used the app, whether it was a helpful addition to their treatment at the study site, and what was perceived as the best and worst things about the study app. The combined response set from these questions provided a rich source of insight on how participants used the app and what sources of benefits they perceived, with approximately 40% of participants reported making some change in their behavior or lifestyle as a result of using the MFVB tool and approximately 30% mentioned perceived benefits to their wellbeing.

#### How the app was used

Participants were asked to “Briefly explain how you’ve used the mental fitness app and how it has (or has not) been helpful to you”. Participants reported a variety of uses, with some describing the app as akin to a therapeutic self-reflection exercise, while others found it challenging to use consistently due to issues with notifications and varying levels of engagement. Many noted that the app helped them track and reflect on their mental health, recognize patterns, and provided a structured daily check-in. However, some participants questioned the accuracy of the MFVB assessment, as it did not always align with their self-assessment. The time constraint for recordings (30 seconds) was a concern for several participants, who desired a longer recording option.

#### Complement to clinical treatment

Although neither the app nor the study was integrated with treatment at the study site, participants were also asked to “please briefly explain how the mental fitness app has (or has not) been a helpful addition to your treatment at CBI”. This question was intended to reveal whether participants would develop their own ways to complement their treatment and how it might be most helpful in a treatment context.

Feedback themes partially overlapped with responses to the previous question, as several participants noted that it helped them reflect, self-assess, and maintain a routine. Some mentioned that it offered a moment for self-reflection and set a positive tone for the day or helped them track their emotions when they didn’t have therapy appointments. However, others found it challenging to remember daily use or believed that it didn’t significantly contribute to their treatment. A few participants mentioned discrepancies in the MFVB score, which didn’t always align with their self-assessment. While some found it beneficial for voicing their thoughts and feelings, others felt it did not align with the specific needs of their therapy.

#### Best and worst things

Participants valued the app’s convenience and ease of use, highlighting its quickness and ability to track emotions over time. They appreciated the score system for visualizing feelings and patterns. The prompts, speech-to-text journaling, and the reminder for positivity were well-received. The app served as a bridge between therapy sessions, offering structured daily check-ins and journaling. Participants also found the tips and score breakdowns into the 8 vocal features helpful, providing a consistent and reassuring external perspective on their emotions.

On the other hand, participants encountered various challenges and limitations with the app, including difficulties in consistently remembering to use it, technical issues such as app glitches and notifications, and the need to find a quiet place to record. The fixed 30-second recording time posed a constraint, leading to desires for greater self-expression flexibility. Repetitive prompts caused some users to lose interest, while doubts about the accuracy of mood interpretation based on voice recordings emerged. Some concerns were raised about privacy and potential data mining.

### Primary endpoint: subgroup analyses

#### Gender and age

RR patterns using the time-weighted MFVB approach for total symptom severity appear to indicate better performance in males vs. females as the RR Ratio is higher (5.00 vs. 1.94). However, confidence intervals for males are wide due to relatively fewer participants, and the data does not provide conclusive evidence for a difference. Still, differences in vocal changes that may impact MFVB effectiveness cannot be ruled out, because gender differences in depressive symptom profiles have been described for other non-voice related behaviors (62).

Analysis by age was hampered by the fact that the middle age group had only one MFVB score in the “Pay Attention” range, and the estimated RR Ratio at 0.61 for this age category has very wide confidence intervals (0.05-7.47, p=0.2276). The younger and older age groups were somewhat better balanced in MFVB scores, and both had RR Ratios consistent with the population at 1.47 and 3.61, the latter being statistically significant.

#### Clinical diagnosis

Subgrouping by clinical diagnosis revealed the most consistent relationship between MFVB score category and RR for depression and stress- and trauma-related conditions, although neither reached statistical significance. For conditions in the anxiety and “other” categories, RR for MFVB scores of “Excellent” and “Good” were as expected but showed little increase in the “Pay Attention” range. Note that the analysis reported here using total M3 score stratified by clinical diagnosis differs from the previously reported RR analysis by mental health symptom type (Table 5), where RR estimates were reported on the entire analysis cohort (all clinical diagnoses combined) separated by symptom type (depression, anxiety, PTSD and bipolar). That said, both analyses indicate best performance in depression and stress- and trauma-related categories, followed by anxiety. While such correspondence in MFVB performance measured either as a function of clinical diagnosis or symptom type appears reasonable on grounds that patients with a given diagnosis would be expected to have the strongest symptoms within that condition, other factors that may impact MFVB effectiveness such as engagement with study procedures, proper use of the study app, overall severity of symptoms or ability to provide accurate self-reported information could differ between these groups.

#### Engagement level

Subgrouping by engagement level reveals that for the high engagement group the RR estimates are considerably more pronounced than for the overall study sample or any other subgrouping variable. This subgroup has an RR Ratio of 8.50 (2.31-31.25, p=0.0013). For the medium engagement subgroup a less pronounced RR Ratio of 1.71 (0.90-3.26, p=0.0996) is observed, and finally in the low engagement subgroup low RR Ratio is observed but with confidence intervals that include 1. This subgroup is characterized by higher symptom burden and fewer M3 scores in the normal-to-mild range, posing limitations on accurate RR estimation.

**Table 8.**
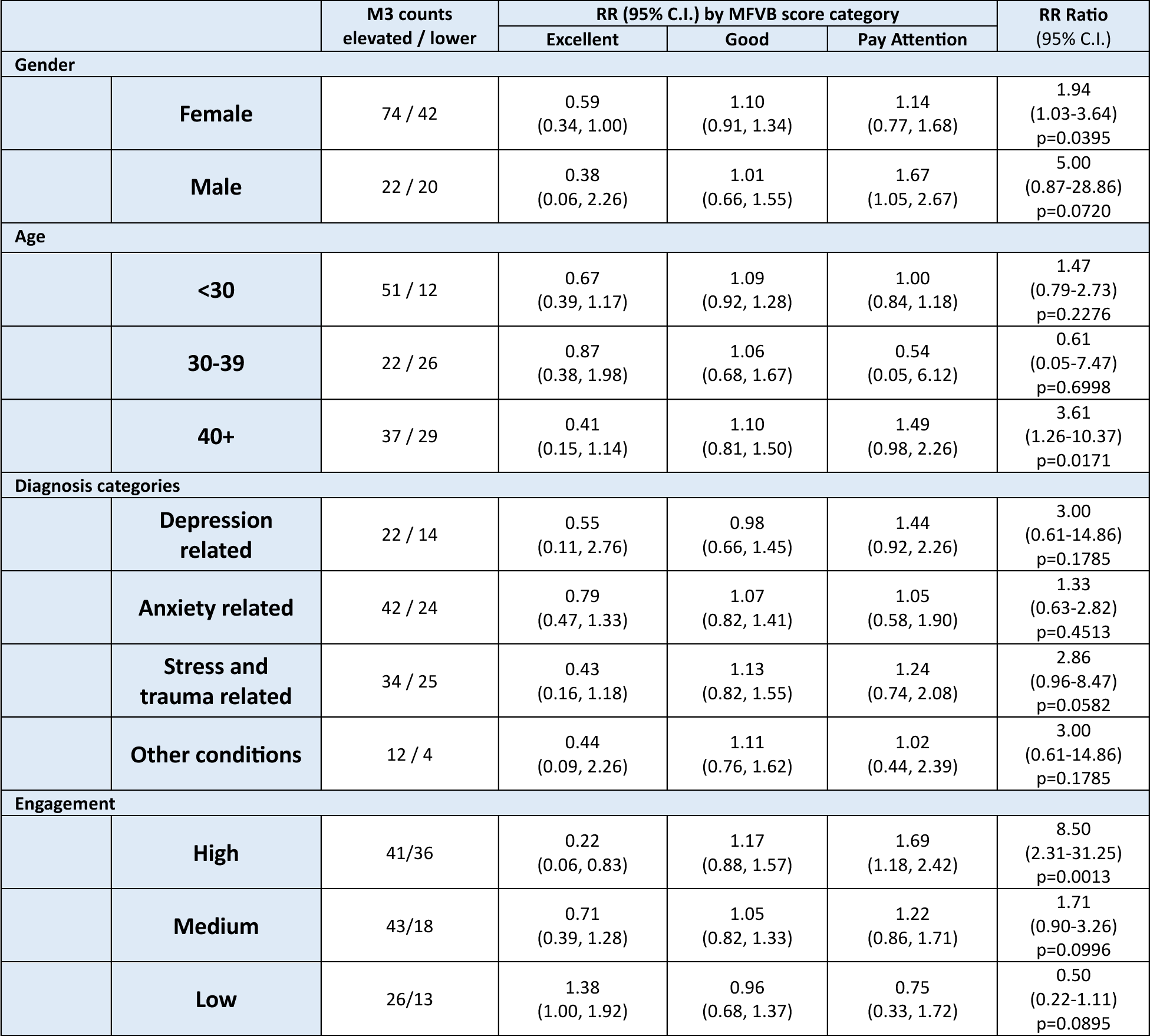
Relative risk for MFVB score categories by selected subgrouping variables. These results are computed with the time-weighted MFVB method (see main text and Fig. 2).

|  |  | M3 counts<br>elevated / lower | RR (95% C.I.) by MFVB score category |  |  | RR Ratio<br>(95% C.I.) |
| --- | --- | --- | --- | --- | --- | --- |
|  |  |  | Excellent | Good | Pay Attention |  |
| Gender |  |  |  |  |  |  |
|  | Female | 74 / 42 | 0.59<br>(0.34, 1.00) | 1.10<br>(0.91, 1.34) | 1.14<br>(0.77, 1.68) | 1.94<br>(1.03-3.64)<br>p=0.0395 |
|  | Male | 22 / 20 | 0.38<br>(0.06, 2.26) | 1.01<br>(0.66, 1.55) | 1.67<br>(1.05, 2.67) | 5.00<br>(0.87-28.86)<br>p=0.0720 |
| Age |  |  |  |  |  |  |
|  | <30 | 51 / 12 | 0.67<br>(0.39, 1.17) | 1.09<br>(0.92, 1.28) | 1.00<br>(0.84, 1.18) | 1.47<br>(0.79-2.73)<br>p=0.2276 |
|  | 30-39 | 22 / 26 | 0.87<br>(0.38, 1.98) | 1.06<br>(0.68, 1.67) | 0.54<br>(0.05, 6.12) | 0.61<br>(0.05-7.47)<br>p=0.6998 |
|  | 40+ | 37 / 29 | 0.41<br>(0.15, 1.14) | 1.10<br>(0.81, 1.50) | 1.49<br>(0.98, 2.26) | 3.61<br>(1.26-10.37)<br>p=0.0171 |
| Diagnosis categories |  |  |  |  |  |  |
|  | Depression<br>related | 22 / 14 | 0.55<br>(0.11, 2.76) | 0.98<br>(0.66, 1.45) | 1.44<br>(0.92, 2.26) | 3.00<br>(0.61-14.86)<br>p=0.1785 |
|  | Anxiety related | 42 / 24 | 0.79<br>(0.47, 1.33) | 1.07<br>(0.82, 1.41) | 1.05<br>(0.58, 1.90) | 1.33<br>(0.63-2.82)<br>p=0.4513 |
|  | Stress and<br>trauma related | 34 / 25 | 0.43<br>(0.16, 1.18) | 1.13<br>(0.82, 1.55) | 1.24<br>(0.74, 2.08) | 2.86<br>(0.96-8.47)<br>p=0.0582 |
|  | Other conditions | 12 / 4 | 0.44<br>(0.09, 2.26) | 1.11<br>(0.76, 1.62) | 1.02<br>(0.44, 2.39) | 3.00<br>(0.61-14.86)<br>p=0.1785 |
| Engagement |  |  |  |  |  |  |
|  | High | 41/36 | 0.22<br>(0.06, 0.83) | 1.17<br>(0.88, 1.57) | 1.69<br>(1.18, 2.42) | 8.50<br>(2.31-31.25)<br>p=0.0013 |
|  | Medium | 43/18 | 0.71<br>(0.39, 1.28) | 1.05<br>(0.82, 1.33) | 1.22<br>(0.86, 1.71) | 1.71<br>(0.90-3.26)<br>p=0.0996 |
|  | Low | 26/13 | 1.38<br>(1.00, 1.92) | 0.96<br>(0.68, 1.37) | 0.75<br>(0.33, 1.72) | 0.50<br>(0.22-1.11)<br>p=0.0895 |

To investigate whether the marked RR improvements with increasing engagement were due to having more MFVB scores available for association with M3 results during the 2-week aggregation window or other factors, RR estimates and RR Ratios were calculated as function of engagement group using the closest-MFVB approach, which uses a single MFVB score only with each M3 result. This approach should diminish the potential advantage of more MFVB scores in the high engagement subgroup. Indeed, this yielded lower RR Ratios of 2.5, 1.3, and 0.90 for the high, medium and low engagement subgroups, suggesting the important contribution of aggregating MFVB results over time.

To further balance the comparison between engagement groups, the closest-MFVB analysis was conducted again using the onboarding M3 only, as the low engagement group was comprised of a large proportion of non-completers which did not provide responses to the offboarding survey at the end of week 4. Isolating the analysis to the onboarding M3 only (which was provided by all participants) will further equalize the data available for association of MFVB results to M3 assessments. This yielded an additional decrease in RR Ratios to 1.5, 1.2, and 0.79 for the high, medium, and low engagement subgroups, respectively. This further solidifies the notion that aggregation of MFVB results over time is the main driver for improving effectiveness of vocal analysis.

## DISCUSSION

### MFVB scores stratify participants into mental symptom severity groups

This study demonstrates the practical deployment of vocal biomarkers in a prospective cohort study, providing participants with real-time insights into their mental health symptom severity. Despite advancements in identifying voice-related features and predictive analytics, translating these approaches into practical technology had remained a challenge, and relatively few published works combine all the aspects tested in our study. To our knowledge, this is the first demonstration of a vocal biomarker tool in any health or wellness application that has delivered meaningful health-related information to actual users in real-time, with a statistically significant level of efficacy.

One recent smartphone app-based speech analysis approach using semantic information for depression and anxiety screening in a mostly older population and compared 5-min voice recordings with PHQ-8 and GAD-7 as reference as a validation study and showed good performance (63). The analysis approach differed from ours in not using acoustic features and providing only binary classifications, without feedback to participants, making it less suited as a personalized mental wellbeing tool. Each session required 5 minutes of voice recording time, significantly more than is required for purely acoustic and prosodic feature extraction. A uniquely long-term (6-month) study in bipolar patients tested whether a machine-learning based vocal biomarker approach could make use of repeated observations over time to identify euthymia, hypomania, and manic episodes in participants via weekly telephone interactions (64), and reported good performance for differentiating hypomania and depression from euthymia. As we found in our study, continuous observations over time proved essential, especially for bipolar disorder, characterized by fluctuating mood over time.

Our method, averaging normalized values of eight vocal features from a 30-second speech sample (see Supplemental Material), facilitates measurement of moderate-to-severe (elevated) mental health symptom severity in terms of Relative Risk (RR), laying a foundation for application-specific products that offer objective and quantitative mental fitness information. RR offers comparative understanding of the likelihood of elevated symptom severities being present within a range of Mental Fitness Vocal Biomarker (MFVB) scores compared to an overall population average.

The use of RR instead of accuracy or sensitivity and specificity, typically employed as outcome measures in vocal biomarker studies, is appropriate given the likely intended uses, which would not primarily involve the identification or differentiation of mental health conditions. The underlying vocal features were selected based on reported correlations with depression but do not have well-supported threshold values that separate clinical categories. Interpretation of MFVB score in terms of RR avoids potentially incorrect interpretation by users that a high (or low) MFVB score rules out (or in) the presence of a mental health condition, whereas the risk concept more appropriately conveys a change in likelihood of underlying symptoms.

We found that MFVB scores also stratified symptom severities within subdomains of depression, anxiety, and PTSD. The RR Ratio was greatest for depression, perhaps explained by the fact the underlying vocal features were primarily selected based on pre-existing evidence for that condition. What is unique about the current study is the transdiagnostic participant sample, applying a uniform vocal processing approach to demonstrate MFVB efficacy for multiple and potentially comorbid mental health conditions. Another differentiating feature of our work is the inclusion of diagnosed individuals only, without healthy controls, aiming to differentiate symptom severity rather than diagnosis status.

### Optimal MFVB performance requires frequent observations over time

We employed two methods for linking MFVB scores to M3 results: one associating a single MFVB score closest in time to the M3, and the other averaging all MFVB results within 2 weeks before or after the M3 (applying weights to diminish the contributions of more distant MFVB scores). Both approaches affirm that MFVB scores provide valuable information about symptom severity. Notably, the time-weighted method exhibits superior risk stratification ability, capitalizing on the accumulation of vocal information over time and aligning more closely with the M3 time window. This distinction, pointing toward the significance of longitudinal observations, is not merely an accidental statistical outcome but an expected behavior. Analogous to understanding a region’s climate, reliable insights into overall mental fitness—the backdrop against which day-to-day mood fluctuations occur—require frequent observation over time. Vocal biomarker tools are inherently well position to gather and refine insights over time, emphasizing the need for future studies and product designs to leverage this intrinsic quality.

Our results confirmed the utility of frequent measurement over time in another way, by revealing significantly enhanced RR estimates in participants that were more highly engaged with the study app. We were able to determine that the enhancement was primarily the result of more frequent “observation” (voice sample recordings). This is good news, because it implies that improvements in MFVB performance can be achieved by boosting engagement for users that would otherwise have low engagement levels. Achieving this goal will require research into strategies that can increase engagement, perhaps tailored to specific user characteristics. An alternative option is to develop passive voice monitoring tools that can continuously observe the user’s voice in the background, once consent has been provided. This would remove the engagement element completely and allow a maximum of observation to occur for all users.

### Participant engagement

As we found above, achieving and sustaining high engagement is essential for many health apps, but often proves challenging in practice. Although the duration of this study is relatively brief, the observed engagement levels are promising. This is evident from the proportion of participants engaged, which combined the mid and high engagement groups, reaching 71%, and the persistence of that engagement, with an observed 87% retention in week 4 averaged across these two groups.

These metrics compare favorably to most other app-based or mHealth studies (65,66). Study design factors that have previously been found to promote retention that were present in this study are the recruitment method (study invitation sent from participant’s clinical treatment organization), financial compensation for participation, and having a relevant condition to the app design (mental health concerns). Perhaps the most important success factor was delivering real-time useful information (MFVB scores) back to users through the study app. The combination of insight and engagement that this approach fosters could be a natural way to enhance the perceived value and success of other digital therapeutic products.

Although participants were offered a financial incentive of $15 per week to encourage app usage of 4 or more times weekly, the incentive did not drive high levels of engagement among all participants. The proportion of participants receiving gift cards in each week was, not surprisingly, correlated with engagement level. That proportion was about 80% in the high engagement group, about 40% in the medium engagement group, and near zero in the low engagement group. Further, a potential small positive effect on app usage in weeks where a gift card was *not* received for the prior week was observed for participants in the low and medium engagement groups.

Engagement was negatively correlated with mental health symptom severity, both for total symptoms and within symptom categories. In particular, the prevalence of elevated depressive symptoms was relatively high in the low engagement subgroup, although the same trends were observed for bipolar, PTSD, and anxiety. Individuals experiencing more severe symptoms may find it challenging to engage consistently with the study app due to the potential impact of their mental health condition on motivation, energy levels, and overall well-being. Higher symptom severity often correlates with increased psychological distress and reduced functional capacity, making it difficult for individuals to sustain regular and active participation in the study. Therefore, the observed trend of lower engagement among those with higher symptom severity aligns with expectations based on the potential influence of mental health symptoms on individuals’ ability and willingness to engage in additional activities. Targeted and higher-intensity engagement and retention strategies may therefore be beneficial for participants with high symptom burden in future studies.

Engagement was positively correlated with age, an effect that has been observed previously in digital health app studies (65,66); although it is contrary to conventional expectations that it is more difficult to engage older populations with digital tools. In this study, age was also negatively correlated with symptom severity; it is therefore possible that age and symptom severity both reinforced the observed engagement trends.

### Study app perception differed by engagement groups

Comments made by participants from the high engagement group indicate a strong commitment to the Mental Fitness app, appreciating the daily check-ins and addition to their daily routines. They found the app beneficial for tracking their mental wellbeing and for self-expression. Participants in the medium engagement group displayed more mixed sentiments, with concerns about notifications and scoring accuracy affecting their overall engagement. The low engagement group participants exhibited the least enthusiasm in their feedback, with limited app use due to forgetfulness or perceived inconvenience, although some participants still acknowledged potential benefits from more frequent use. This seems consistent with the 53% level of agreement stated by the low engagement group in the end of study feedback, when asked whether they intend or would like to keep using the app to track their future wellbeing – a number well above their actual level of use (even the subset of low engaged users that completed the feedback).

Perceived accuracy of the MFVB scores was expected to have an impact on engagement, but this was not evident from the feedback. The low-engagement group made no comments about lack of accuracy, while high- and medium-engagement groups commented similarly on perceived accuracy (or lack thereof) of the MFVB scores. Interestingly, Table 7 indicates that the medium engagement group has the lowest level of agreement with the statement that the MFVB scores are helpful in understanding how they are doing, even less than the low engagement group. This appears consistent with the summarized free response feedback described above, and the perception of limited benefits for personal insight may be a key factor holding these participants back from more consistent app use.

When considering whether the MFVB tool is a beneficial addition to mental health treatment, this study has revealed how digital health apps may provide complementary benefits to therapy sessions. Participants that frequently used the app say that they generally found the app to be a valuable addition to their treatment as a tool for self-reflection, providing reminders to relax and a way to practice the skills they worked on in therapy. It helped them focus on positive thoughts and stay in a mental health routine. Less engaged users also appreciate the tool as a platform to voice thoughts and feelings but do not perceive it as contributing to their treatment as much, either because they do not use it as frequently or because they view it more as a mental exercise.

### Limitations

#### Study population

The main limitations of the present study are related to the study population, which was an outpatient psychiatric sample with limited demographic diversity in race and ethnicity. Although a large age range was represented (16-80), most participants were young adults to early middle age. All were native or fluent English speakers and clients of a mental health care provider based in Pittsburgh, PA. On the other hand, the MFVB scoring algorithm was developed based on vocal feature analysis from a large Indian outpatient population speaking 5 different Indian languages, which would suggest that our results are not particularly sensitive to linguistic, geographic, or cultural differences. Reliance on acoustic features vs. linguistic analysis is likely a major contributing factor to this robustness. Given the marked difference in the development and validation cohorts, we hypothesize that many of the findings in this report would generalize to other populations as well.

The clinical population included in this study may exhibit certain characteristics that would be different in a more general population. For example, the study participants are actively engaged in mental health treatment and volunteered for the study, which suggests a certain level of motivation to engage with tools intended for mental fitness tracking. They were also financially incentivized to use the app. Other populations may exhibit different interests and uses for the tool studied here. We also found that those participants with more severe symptoms, in particular those related to depression, were on average less engaged relative to the participants with milder symptoms. These participants with milder symptoms may share many characteristics with a portion of the general population, given estimated levels of depression and anxiety in United States in the 30-35% range (67).

#### Limitations in study app functionality and user interface

Our findings regarding engagement levels and participant feedback are influenced both by the MFVB scoring algorithm and the overall app experience. Because our MFVB tool was presented as a voice-to-text journaling app, many participants reported valuing the combination of recording thoughts and moods, self-reflection, and tracking their wellbeing through the MFVB scores. Users also valued other aspects including notifications to help build habits, tips for healthy activities and behaviors, etc. This points to the need to consider the entire product experience and user journey in the context of digital health products like vocal biomarker assessment. Because this app was built primarily as demonstration and limited research tool, the positive reception and usage levels would likely further increase in an application more optimized for user experience and customization.

#### Lack of diagnostic specificity

Our results demonstrate a general ability of MFVB score categories to differentiate mental health symptom severity levels, but do not distinguish what type of symptoms these may be (e.g., depression, anxiety, PTSD, or bipolar). The eight acoustic and prosodic features that underlie the MFVB score algorithm (see Supplemental Material) capture enough voice acoustic changes that accompany the presence of these conditions that the resulting MFVB score is generally responsive transdiagnostically, serving as a kind of “mental wellbeing thermometer”. It is possible that the combined pattern of changes among the 8 vocal features could differentiate among these mental illnesses, although no existing work is known to the authors that has demonstrated such capability for the disorders included in this work. In a vocal biomarker study of people with depression, bipolar, schizophrenia and healthy controls, it was found that it was possible to train classification models to differentiate the disordered participants from healthy controls, but not depression from bipolar (68). Schizophrenia could be differentiated from depression and bipolar but falls outside the scope of common mood disorders that is being targeted with our Mental Fitness approach.

### Potential applications and future work

#### Application examples

The most obvious application might be the one in which this study was conducted: mental health treatment, where patients can benefit from objective insights into their mental wellbeing over time. The information may also be useful to providers, allowing a complementary source of insight and, through a non-fatiguing assessment at relatively greater time resolution, filling temporal gaps between treatment sessions. Additionally, even users who are not actively undergoing mental health treatment may benefit from monitoring their mental fitness. Further exploration could involve integrating MFVB scores into workplace wellness programs, offering employers a tool to support employee mental health. This technology could be integrated into digital health platforms and wearables, extending the reach of these offerings and offering a more holistic view of well-being that includes mental aspects as well as traditional fitness measures such as step count, sleep, and cardiovascular metrics. Another avenue for exploration is the development of prognostic outcomes that could be used, for example, in primary care to identify those individuals most likely to be diagnosed with a mental disorder upon referral to a specialist. The potential extends to services targeting adolescents and college-age populations, considering that approximately 20% of our study participants belonged to this age range, with results consistent with the overall study cohort.

#### Vocal analysis in the background

While we have shown that the active journaling component of the MFVB approach is valued by many users, some did not use it and others forgot to use the app consistently or found it difficult to incorporate into their daily life. Sonde Health has already developed passive measurement capabilities to analyze user voice in the background, which allows the MFVB results to be provided without active user engagement. The technology runs on edge devices rather than hosted on cloud platforms, eliminating the need for recording or transmitting voice data. This not only addresses privacy concerns but also aligns with evolving data protection standards. Passive monitoring also includes user voice identification to ensure that analysis pertains solely to the intended user, further enhancing confidentiality of the mental fitness solutions. Future studies will incorporate such passive measurement capabilities to address engagement and privacy challenges.

#### Optimization of processing and scoring algorithms

This study did not seek to optimize MFVB scoring algorithms to maximize risk stratification abilities; therefore, a relatively simple combination of pre-selected voice acoustic features was used for scoring. The current algorithm’s simplicity likely contributed to its robustness from the training data set (outpatient clinics across regions in India) to this validation cohort. Additionally, this study’s positive results are an encouraging foundation for future algorithm development, including machine learning or artificial intelligence approaches, to improve alignment of MFVB scores and mental health symptoms. The study app also did not include any voice sample quality control mechanisms to reject recordings with high background noise, which, if present, could negatively impact score accuracy. Including such control mechanisms may further enhance MFVB score performance.

#### Personalized change detection

The MFVB scoring algorithm in the study app, derived from cross-sectional data, assigns scores and category labels based on population vocal feature distributions, limiting direct applicability to individual users due to voice variations and smartphone device differences across users. While participants found tracking MFVB changes over time useful, determining the clinical significance of individual score changes was not possible. Longitudinal studies, assessing concurrent symptom and vocal changes, are needed. The study’s 4-week duration was inadequate to capture meaningful mental condition changes for most participants; for instance, 74% of participants did not change in symptom severity category, perhaps because most participants had been in treatment for an extended period of time already (66% >1 year, Table 2). A 12-week extension phase of the study aims to assess individual changes and potentially inform personalized scoring algorithms and will be reported in future communications.

#### Impact on mental wellbeing

Given the positive experiences participants reported with the MFVB tool, it would be of interest to study if and how mental fitness tracking might improve mental wellbeing, akin to the well-establish impact that step-count monitoring has on physical activity and its attendant physical health benefits (40). Via the study feedback, approximately 40% of participants reported making some change in their behavior or lifestyle as a result of using the MFVB tool and approximately 30% mentioned perceived benefits to their wellbeing – while neither of these themes were specifically included as questions in the survey. These are remarkable statistics for a basic smartphone app that merits follow-up studies designed to measure the potential impact of an MFVB tool as health intervention.

## CONCLUSIONS

We have demonstrated that a Mental Fitness Vocal Biomarker (MFVB) scoring algorithm, using pre-selected vocal features reported in the literature, incorporated into a smartphone voice journaling application can indicate risk ratios for elevated mental health symptom severity of 1.53 from a single 30-second voice sample. By aggregating recordings over a 2-week period this increases to 2.00, and for highly engaged users up to 8.50. Similar performance was observed within mental symptom categories of depression, anxiety, and PTSD. Over 70% of study participants were consistently engaged, on average using the MFVB tool 3-4 times per week over 4 weeks. 70-80% of participants enjoyed using the tool, understood the results, and found the MFVB scores helpful in understanding how they were doing. Participants mentioned benefits such as creating helpful daily routines, therapeutic benefits from self-expression, and valuing the fact that they could contrast their self-perception with an objective assessment.

While the MFVB tool is not intended to diagnose or treat mental health conditions, these findings illustrate its meaningful potential in personalized wellness tracking, which has so far not yet been able to extend measurement of physical health to mental wellbeing. Its functional appeal, consistent use, and positive reception highlight its significant potential to benefit users. The key insight from this work – aggregating MFVB scores over time and integrating information amid day-to-day mood variations – is an important factor in attaining optimal performance levels. Vocal biomarker-based wellness tools are well-suited for continuous observation, engaging users to cultivate healthy habits and behaviors, ultimately contributing to improved wellbeing.

## Supporting information

Supplemental Methods Section

## Data Availability

All data except participant voice recordings produced in the present study are available upon reasonable request to the authors and execution of a data use agreement with Sonde Health

## ABBREVIATIONS

MFVB: Mental Fitness Vocal Biomarker
RR: Relative Risk

## ACKNOWLEDGMENTS

The authors wish to acknowledge Krushnakant Mardikar and Mrityunjay Samanta from gslab for significant contributions to data analysis and signal processing methods. The pilot phase of the study at St Joseph’s Healthcare Hamilton was partially supported through the Mitacs Accelerate International – from Canada program.

## Footnotes

1 The RR Ratio as defined in this paragraph is equivalent to the risk ratio between the “Pay Attention” and “Excellent” categories, as the reference group used in the <u>relative</u> risk calculation factors out when calculating this ratio. However, in some subgroup analyses the contingency tables include zeroes that would cause computational problems, and in those cases a value of 0.5 is added to all cells. This is done separately for the calculation of relative risk by MFVB category and risk ratio between the aforementioned MFVB categories. This results in a few instances where the ratio of reported relative risks values for “Pay Attention” vs. “Excellent” appear numerically different from the reported RR Ratio values (which are calculated with a different contingency table).

