## Supplemental Methods Section for "Validating the efficacy and value proposition of Mental Fitness Vocal Biomarkers in a psychiatric population: prospective cohort study"

### SUPPLEMENTAL MATERIAL

#### Vocal biomarker selection

A literature survey was conducted to identify a group of well supported vocal features relevant to mental health conditions, with a focus on depression. No formal selection criteria were employed, although preference was given to those features that had more substantial evidence for correlation with mental conditions; interpretability of the features was also considered. Finally, preference was given to features where the implementation could be validated with existing vocal analysis tools, in particular OpenSMILE and Praat. This resulted in eight features being selected, summarized in Table 1 in the main article, which includes feature value distributions from an internal reference dataset and voice samples obtained in this study (address in the Results section).

The eight pre-selected vocal features together represent different aspects of voice and speech production; supporting citations are provided in the main article. Vocal cord control is represented by the glottal measures jitter and shimmer, which measure cycle-to-cycle variations in pitch and amplitude during phonation. These variations have been shown to increase due to reduced motor control in individuals with mental health conditions.

Supra-segmental features related to intonation are captured by pitch variability and energy variability. These features measure the amount of deliberate voice modulation during speech and are among the more recognizable changes that occur in depression (flat affect).

Measures of vocal tract coordination are included as vowel space and phonation duration. Vowel space is a metric that estimates the degree of separation in the first two formant frequencies across the entire set of speech vowels. These frequencies are determined by precise coordination of the articulators and can be impacted by reduced muscular control or psychomotor retardation. Phonation duration measures the average time from onset to offset of glottal vibration (phonation), which is similarly a measure of muscular control.

Prosodic features are included as speech rate and pause duration, which are indicators of psychomotor retardation and speech hesitations, both of which have been linked to depression. Note that these metrics are measured from acoustic analyses, not a count of words based on automatic speech recognition. These features are also implicated in mild cognitive impairment and dementia, which may provide some sensitivity to these conditions.

#### Signal processing considerations

The features described above were extracted using standard speech processing methods and where possible were validated against OpenSMILE and Praat. Feature values were computed in overlapping 30-ms frames, and summarized over the entire recording using means, medians or interquartile ranges as applicable. Only active speech portions of the recording were considered, by using an energy-based Voice Activity Detection (VAD) algorithm. No other pre-processing was performed on the recordings.

#### Mental Fitness algorithm

To simplify interpretation of vocal feature results, a composite score was developed based on the 8 features described above. Because the relative efficacy of these features for mental health assessment has not been previously studied, a simple unweighted averaging procedure was used to compute a "Mental Fitness Vocal Biomarker" (MFVB) score. Before averaging, each feature value was transformed to a normalized value on a scale of 0-100 meant to represent the distribution limits of each feature value. Existing datasets acquired prior to the present study, described below, were used as a guide for these feature value distributions, which are also summarized in Table 1. All vocal features were arranged in the averaging approach so that increasing values correspond to reductions in likelihood of mental health symptoms, so that the resulting MFVB score has the straightforward interpretation that higher values indicate higher mental fitness, and vice versa, on a possible range of 0-100.

Reference vocal feature distributions were based on a vocal biomarker dataset collected as part of a different study from over 50,000 volunteers between August 2018 and January 2020 involving over 20 hospitals in India. All research subjects provided informed consent to a clinical research coordinator at the site of their outpatient visit and were then guided through a series of 4-6 vocal tasks and questionnaires while their responses and voice samples were recorded directly on a smartphone. Vocal responses were recorded primarily in the following languages: Hindi, Marathi, Telugu, Malayalam, and Kannada. Additional demographic and health condition diagnoses metadata were collected by the clinical research coordinator from the medical charts at the time of the visit and entered into a separate system. These data were then transferred and stored securely on a remote server for processing.

From this digital biobank, a subset of data comprising about 20,000 participants was selected after voice sample quality control implemented through a standardized process with trained staff. This subset included approximately equal males and females and was distributed similarly across 4 age groups (18-29, 30-49, 50-64, and 65+). PHQ-9 responses were collected on all participants to study depression symptoms, indicating about 10% of participants had moderate or more severe symptoms (PHQ-9>=10), although <2% of participants had a history of depression diagnosis. Most prevalent diagnosed conditions in this data subset included maternal health (pre- and post-partum), hypertension, coronary artery disease, asthma, hypothyroidism, and migraine (a combined 56% of participants). Healthy volunteers were also included (5% of participants). Vocal feature value distributions were computed from 30 second free speech recordings and used as reference for the normalization procedure described above (see Table 1).

#### Mental Fitness study app

The study used the Sonde Mental Fitness app, available for iOS and Android smartphones. The app is designed to promote voice journaling and incorporates two key functionalities: transcription (provided by Amazon Web Services, AWS) and Mental Fitness scoring, as described above. The combination of these functionalities is intended to promote the potential mutually reinforcing benefits of journaling and MFVB biomarker feedback. The app is intended primarily as a proof-of-concept vocal biomarker showcase and research tool, and is not intended to screen, diagnose or treat any mental health condition.

The user flow through the voice journaling portion of the app includes a mood check-in, recording of context/ reason for mood, a 30-second voice recording, followed by the MFVB score and its individual feature value results. The transcription of the voice journal is available for review in a separate section of the app. Before the voice recording, the user receives a randomized prompt that is intended to give suggestions for what to say and promote self-reflection (e.g., "How are you feeling today?", "Describe things that make you happy", "What are you looking forward to this week", "What you are you avoiding dealing with?"). The user can request a different random prompt or use no prompt to talk about their own topics. Recording duration is fixed to 30 seconds. Users are reminded to seek a quiet location for to minimize the presence of background noise in the recorded audio.

MFVB score results are displayed on a range of 0-100 and include a qualitative label of either "Pay attention", "Good', or "Excellent" for score ranges of 0-69, 70-79, and 80-100 respectively. These ranges were determined from the same dataset as described above, approximately representing the lower quartile, middle two quartiles, and upper quartile of the Mental Fitness score distribution. These labels were chosen to reinforce the intended use of the tool, namely, to promote personalized insights and self-reflection. Simpler labels such as "low", "medium" and "high" could be misinterpreted as directly applying to the user's mental health status and create unwarranted concerns.

The eight voice feature value results are also provided, each one including a plain language explanation of its interpretation. Finally, the MFVB score and each feature value has a history graph showing the 7 most recent results, allowing the user to track trends over time.

To further promote healthy behaviors, the result screens also include 3 randomly selected "tips" displayed as icons with a brief description, suggesting activities or behaviors that are generally understood to be beneficial for mental wellbeing (e.g., "practice meditation", "connect with friends", "express gratitude", "ask for help"). Each icon is linked to independent third-party online web resources with topical information. The app includes two mechanisms to promote consistent use, including optional daily reminders (app notifications and SMS messages that the user needs to enable) and a streak tracker.

#### Privacy and security

The app and user data are securely hosted on Amazon Web Services (AWS). The hosting infrastructure uses AWS security mechanisms, including but not limited to encryption in transit and at rest, identity and access management controls, and regular security audits. To ensure user privacy, sensitive account information, such as personal identifiers and login credentials, is stored separately from other user data within the AWS environment. The app adheres to industry best practices and regulatory standards related to data privacy, including HIPAA and GDPR. Regular security assessments and audits are conducted to identify and address potential vulnerabilities promptly. Terms of Service and Privacy Policy are provided to users within the app; additional privacy controls and rights are applicable to users who are participants in research studies, as described in the study informed consent information (separate from the study app).
